# Global RNAseq of ocular cells reveals gene dysregulation in both asymptomatic and with Congenital Zika Syndrome infants exposed prenatally to Zika virus

**DOI:** 10.1101/2020.10.20.20214403

**Authors:** Livia Rosa-Fernandes, Amina Bedrat, Maria Luiza B. dos Santos, Ana Pinto, E Lucena, Thiago P. Silva, Rossana C. N. Melo, Giuseppe Palmisano, Claudete Araújo Cardoso, Raquel Hora Barbosa

## Abstract

In 2015, Brazil reported an outbreak identified as Zika virus (ZIKV) infection associated with congenital abnormalities. To date, a total of 86 countries and territories have described evidence of Zika infection and recently the appearance of the African ZIKV lineage in Brazil highlights the risk of a new epidemic. The spectrum of ZIKV infection-induced alterations at both cellular and molecular levels is not completely elucidated. Here, we present for the first time the gene expression responses associated with prenatal ZIKV infection from ocular cells. We applied a recently developed non-invasive method (impression cytology) which use eye cells as a model for ZIKV studies. The ocular profiling revealed significant differences between exposed and control groups, as well as a different pattern in ocular transcripts from Congenital Zika Syndrome (CZS) compared to ZIKV-exposed but asymptomatic infants. Our data showed pathways related to mismatch repair, cancer, and PI3K/AKT/mTOR signaling and genes probably causative or protective in the modulation of ZIKV infection. Ocular cells revealed the effects of ZIKV infection on primordial neuronal cell genes, evidenced by changes in genes associated with embryonic cells. The changes in gene expression support an association with the gestational period of the infection and provide evidence for the resulting clinical and ophthalmological pathologies. Additionally, the findings of cell death- and cancer-associated deregulated genes raise concerns about the early onset of other potential pathologies including the need for tumor surveillance. Our results thus provide direct evidence that infants exposed prenatally to the Zika virus, not only with CZS but also without clinical signs (asymptomatic) express cellular and molecular changes with potential clinical implications.

## Introduction

In 2015, Brazil reported an outbreak identified as Zika virus (ZIKV) infection, transmitted mainly by *Aedes* mosquitoes^1,2^. The transmission of this disease, associated with Guillain-Barré syndrome and microcephaly, has been registered in different regions of the world. To date, a total of 86 countries and territories have described evidence of Zika infection (transmitted by mosquitoes) ^1^ and, more recently -in 2019, the appearance of the African ZIKV lineage in Brazil highlight the risk of a new epidemic^3^.

Congenital Zika Syndrome (CZS) was identified due to the increased incidence of congenital abnormalities associated with ZIKV infection, which include morphological, behavioral, neurological^4,5,6,7^ and ocular impairments^8,9,10,11,12,13^. Notably, not only ZIKV antigens have been detected in ocular tissue samples^14^, but also ZIKV has the potential to survive for long periods in ocular tissue and it is detectable in ocular fluids (e.g. lacrimal fluid)^15,16,17^.

Ocular lesions may result from defects of eye development that may have occurred at different stages of embryogenesis^8,9,10,11,12,13,14^. On the other hand, the eye has been described as a specialized CNS compartment, since the optic nerve and retina extend from the diencephalon during embryonic development, being able to display symptoms of neurodegenerative diseases^18^.

ZIKV infection might yield outcomes that will only be manifested later in life^19^, requiring long-term monitoring of all infants prenatally exposed to ZIKV^20^,^21^. Considering the need for after birth surveillance of infants with possible ZIKV congenital exposure, we have standardized an impression cytology methodology and study protocol for analysis of molecular alterations resulting from ZIKV infection in ocular tissue^22^.

Here we present the first study of global gene expression responses of CZS from ocular samples captured by impression cytology. The ocular profiling revealed significant differences between exposed and controls, as well as a different pattern in ocular transcripts from CZS compared to asymptomatic infants. Congenital ZIKV exposure triggered the regulation of pathways related to mismatch repair, cancer, and PI3K/AKT/mTOR signaling and genes probably causative or protective in the modulation of ZIKV exposure. Altogether, our results enlighten the understanding on the molecular effects of ZIKV exposure during human embryonic development, getting attention to the need for surveillance of all exposed children, even when born without clinical signs.

## Material and Methods

### Ethical statement and clinical aspects

This study was carried out in accordance with the ethical principles taken from the Declaration of Helsinki and written informed consent was obtained from the parents of babies under Institutional Review Board and Ethics Committee of Universidade Federal Fluminense approved protocols (approved protocols register number - CAAE: 79890517.6.0000.5243). Ocular surface samples were obtained from a total of twelve infants diagnosed according to the CZS diagnostic criteria referred to the Pediatric Service of the Hospital Universitário Antonio Pedro, Universidade Federal Fluminense (HUAP-UFF), Brazil. All children have been followed by periodical clinical examinations including neurological and ophthalmological screening. From eight babies exposed to the Zika virus during pregnancy, four presented clinical characteristics corresponding to CZS while four did not present clinical signs at the time of ocular cell collection (asymptomatic infants). Control samples were obtained from four unaffected infants, with no clinical signs and no exposure to ZIKV during gestation.

### Impression cytology and ocular surface cells capture

Ocular surface samples from the infants exposed to ZIKV were obtained through optimized and non-invasive impression cytology method using a hydrophilic nitrocellulose membrane as previously described^22^. Additionally, cytospin preparations were done to enable concentration of single cells in suspension on a microscope slide and morphological evaluation (qualitative and quantitative) of potential cell changes. All procedures were described in detail in our previous report^22^.

### RNAseq and Data analysis

Cell suspensions (9 μL/sample) containing a maximum number of 150 single cells/sample (nuclei) were placed in individual 0.1 mL PCR tubes with 2.5 μl 1 x PBS (excluding calcium and magnesium) and cells were processed through the Single Cell RNAseq Service (SingulOmics/ Novogene). Processing included RNA isolation, cDNA synthesis and amplification, library preparation, and sequencing (10 million *paired-end reads per sample*)^22^. Reads from RNA-seq were subjected to quality trimming using Trim Galore (version 0.5.0) and aligned to human reference genome (GRCh38.93) using STAR (version 2.6.1a; options: --outSAMattrIHstart 0 --outSAMstrandField intronMotif --outFilterIntronMotifs RemoveNoncanonical --alignIntronMin 20 --alignIntronMax 1000000 --outFilterMultimapNmax 1)^23^. Duplicated reads were discovered using Picard tools (version 2.0.1) and removed. Gene annotations (gtf file; version GRCh38.93) were obtained from Ensembl. FPKM values of genes were estimated using cufflinks (version 2.2.1)^24^. Counts of genes were estimated using Ht-Seq (v0.10.0)^25^ and Ensembl gene annotation file (gtf file; v. GRCh38.93). Moreover, genes with less than 5 normalized counts in one or more samples are excluded from the analysis. The statistical analyses were performed in R v 3.5.0. We processed the data using DESeq2 v1.14.1^26^. The reads counts were normalized, the fold change between groups was calculated, and the differentially expressed genes were selected by adjusting |Log2 FC|> 1 and False Discovery rate (FDR) corrected p-value (Benjamini–Hochberg) <0.05. We then analyzed a set of genes (top 5%-fold change) to find enriched Gene Ontology (GO) terms with respect to the complete set of tested genes (8583). We used the Gorilla web service^27^ with the running mode “Two unranked lists of genes” and default P-value threshold (1⍰×⍰10^−3^).

### Quantitative Analysis of transcriptomic Data and Statistical Analysis

Data were processed using Perseus software (v1.4.1.3, Max Planck Institute of Biochemistry, Martinsried, Germany)^28^. Principal component analysis (PCA) was performed as exploratory data analysis approach in all quantified transcripts. One-way analysis of variance (ANOVA) was used for multiple testing while t-tests were performed for the comparison between two groups. For both tests, p-value< 0.05 were considered statistically significant. Regression and correlation analyses were also performed for the obtained results. The correlations between variables were defined by the Pearson (Perseus) coefficients, and p values less than 0.05 were considered significant. Protein-protein interaction networks were constructed using STRING^29^ software with High confidence parameter (0.700). Pathway and disease enrichment analysis were also carried out ToppFun function of ToppGene^30^ suite with FDR correction of 1%.

## Results

### Clinical aspects

Samples were collected from both eyes of infants (eight boys and four girls) with median age of 21 months (Supplemental Table 1). Four infants were ZIKV exposed and presented CZS according to the CZS diagnostic criteria (ZIKV/CZS: four infants with positive PCR for Zika virus in gestation and presence of clinical signs which included ocular abnormalities and microcephaly – ZIKV infection predominantly in the first trimester), four infants were ZIKV exposed (positive PCR for Zika virus during gestation; maternal ZIKV infection occurring in the second and third trimester) but no clinical signs identified at sample collection time (ZIKV asymptomatic infants) and four unaffected and ZIKV unexposed infants (control samples - CTRL with negative PCR and without clinical signs; Figure 1).

**Figure 1.**
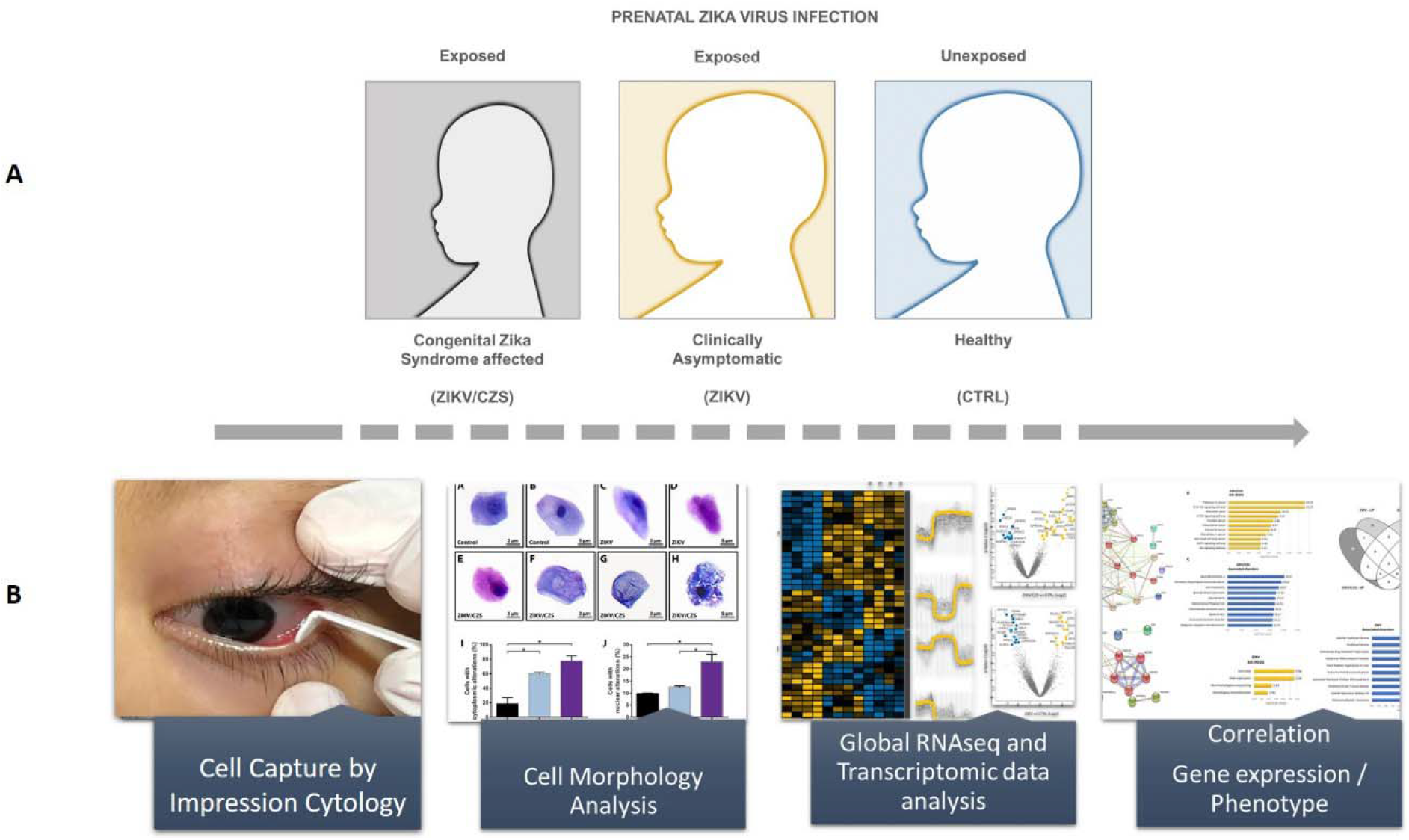
Graphical abstract. **A**. Schematic figure representing the different groups studied - Exposed (CZS and ZIKV) and not exposed (control) prenatally to the Zika virus. **B**. Cell collection procedure and experimental design.

### Ocular Cell surface abnormalities

The morphology of cells collected from exposed and non-exposed infants was evaluated in parallel studies^22,31^. Changes associated with cell degeneration and death were clearly detected in cells from ZIKV-exposed patients, predominantly in CZS, compared to controls and included different levels of keratinization, pyknosis, karyolysis, anucleation and cytoplasmic vacuolization (Figure 2). Quantification of these cytoplasmic and nuclear alterations demonstrated a significant higher alteration in infected infants (Figure 2).

**Figure 2.**
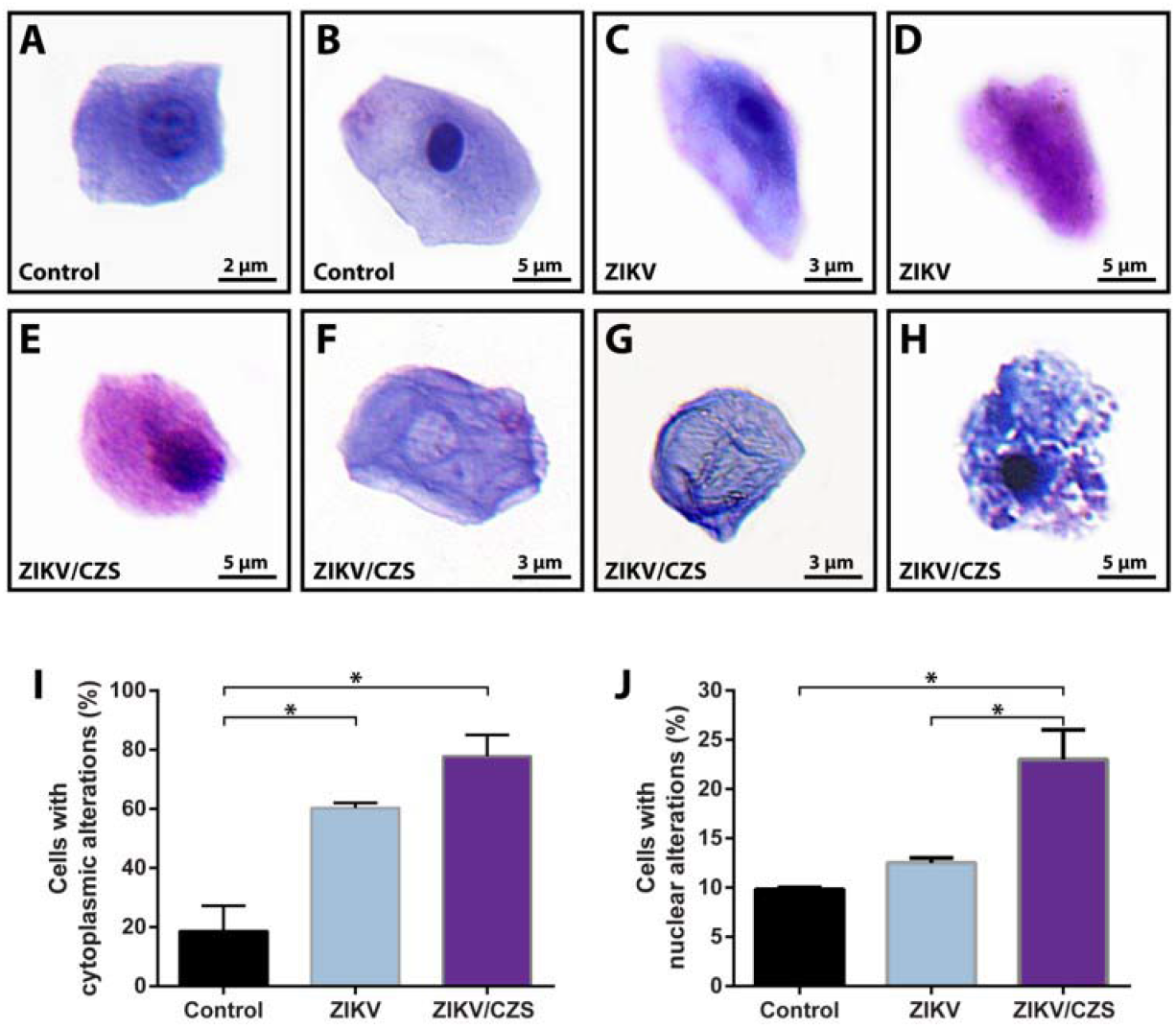
ZIKV infection triggers nuclear and cytoplasmic alterations in human conjunctival cell types. **(A, B)** Representative cells from uninfected group show normal morphology. **(C-I)** Nuclear and cytoplasmic alterations observed in cells collected from ZIKV (infected with no clinical signs) or ZIKV/CZV (diagnosed congenital Zika syndrome) children. Morphological changes were characterized by pyknosis **(C, E, H)**, keratinization **(C, D, E)**, anucleation **(D, G)**, karyolysis **(F)**, vacuolization **(C, H)** and cell fragmentation **(H)**. Quantitative analyses **(I, J)** show increased number of conjunctival cells with morphological changes. Cells were collected by impression cytology, cytocentrifuged and stained with Diff-Quik or toluidine blue. For each group, 100 cells were analyzed in 2 different slides and scored for morphological changes. Data represent mean ± SEM. * *P* < 0.001.

### RNAseq profiling

Transcriptome analysis showed 8582 transcripts quantified in all samples (> 5 reads in all samples; Figure 3A). Principle Component Analysis of these transcripts in all conditions with a non-supervised distribution showed no apparent difference among the samples.

**Figure 3.**
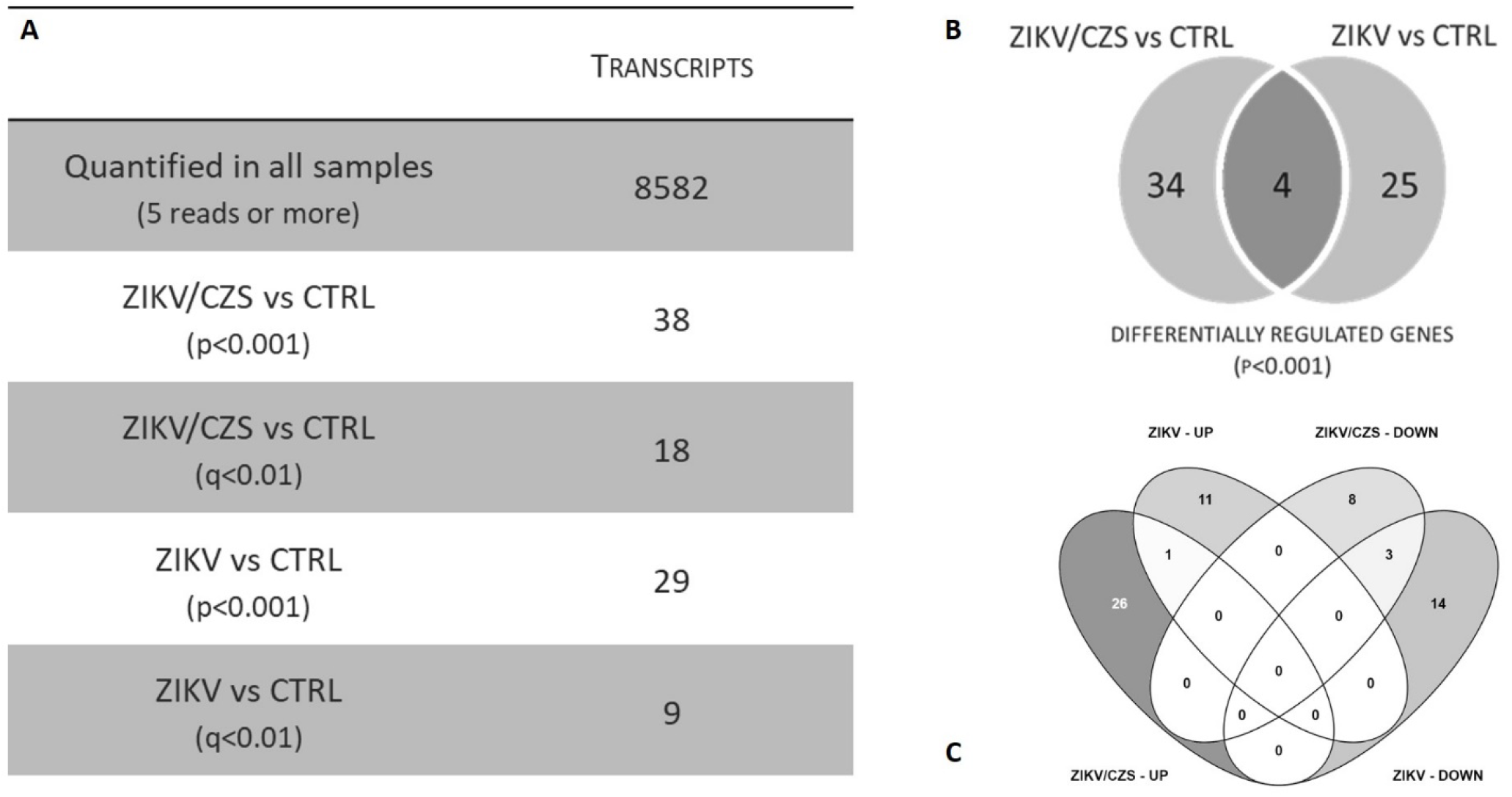
Ocular surface cells gene expression analyses. **A**. Descriptive analysis table of transcripts quantified and regulated. **B**. Venn diagram of DRG (p<0.001) according exposed group. **C**. Differentially expressed gene per exposed group by VENNY analysis.

Ocular cells from exposed infants, ZIKV/CZS and ZIKV, showed 63 differentially expressed genes [p-value<0.001; 27 differentially expressed genes if considering adjusted p-value <0.01], when compared to the unaffected group (CTRL) group. However, the ZIKV exposed groups are heterogeneous within the genes differentially expressed between the two of them. Thus, we proceeded to contrast each of the exposed groups with the unexposed controls (Figure 3C).

A total of 38 genes are differentially expressed in ocular cells from ZIKV/CZS compared to CTRL (Figure 4A and Table 1) while 29 genes are regulated in ZIKV compared to CTRL (Figure 4B and Table 2). Only 4 genes (*SRBD1, MUC21, ALMS1* and *CRACR2A*; p<0.001) are differentially regulated in ocular cells from both exposed groups when compared to CTRL; Figure 3B).

**Figure 4.**
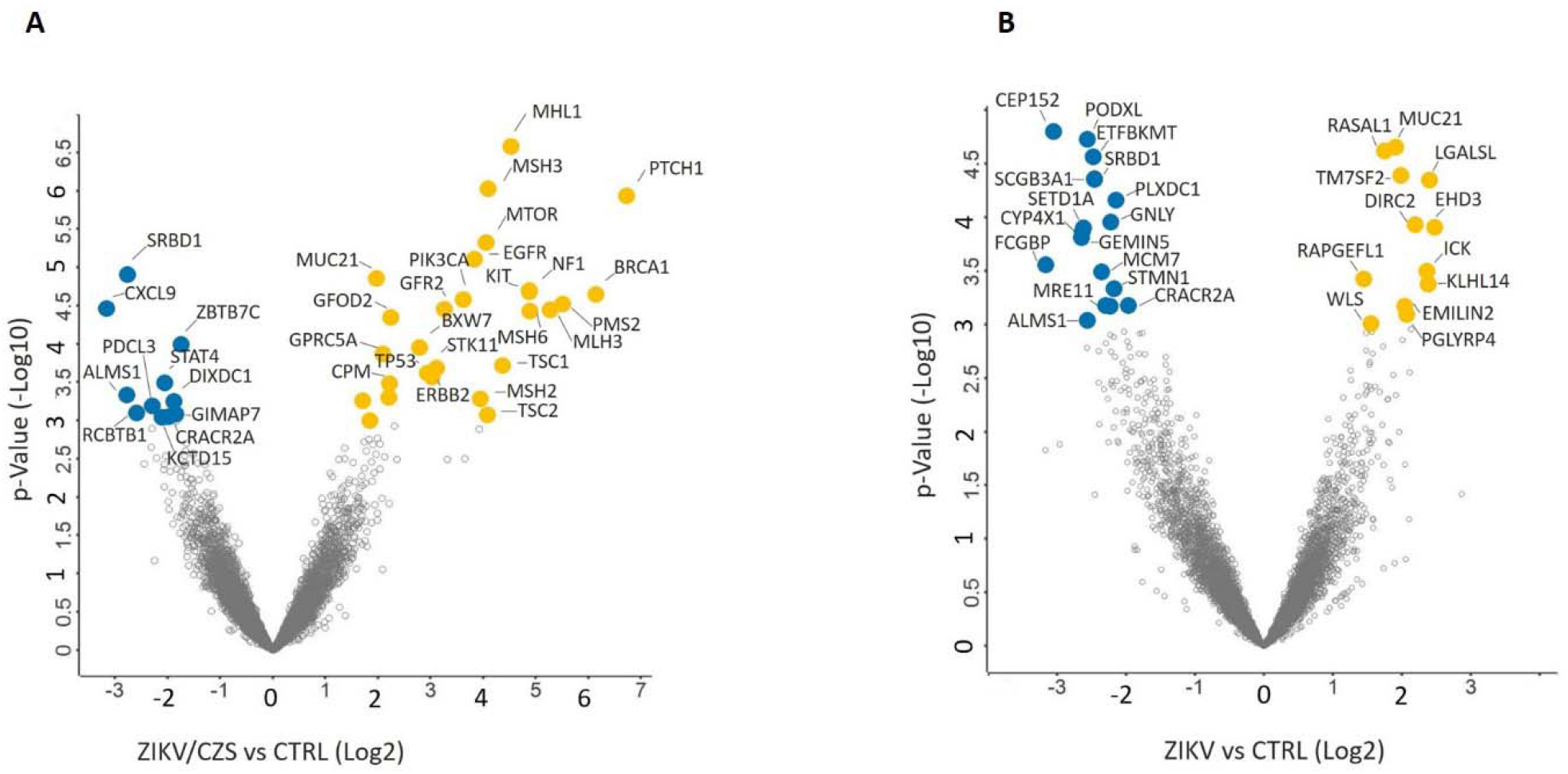
Ocular transcripts differentially expressed comparative analysis A. CZS sample compared to CTRL samples. **B**. ZIKV without clinical signs sample compared to CTRL samples. Volcano plot showed transcripts up-regulated (yellow) and downregulated (blue).

### Comparative transcriptomics

Comparison among ZIKV/CZS, ZIKV and CTRL groups revealed four well-defined hierarchical clusters (Figure 5). The first cluster showed a clear association of genes *ARF3, KLHDC4, HIVEP2, TNFAIP1, ADRM1, GFOD2, TAF13, RAPGEFL1, TM7SF2, LGALSL, ANXA9, TMUB2, PGLYRP4, XPC, EHD3, PROM2, RASAL1* and *MUC21* in most cases downregulated in CTRL infants and upregulated in all ZIKV exposed infants (either CZS and ZIKV, without clinical signs). The second cluster showed association of genes *NUP93, FXYD5, TRIB2, PLXDC1, HDGFL3, SETDB1* and *POLRMT* in most cases downregulated in CTRL and ZIKV/CZS and upregulated in ZIKV exposed infants. The third cluster showed association of genes *MARF1, BTN3A3, FAM102B, OSER1-DT, BICRAL* and *PLA2G4A* in most cases downregulated in ZIKV/CZS and upregulated in CTRL and ZIKV exposed infants. The fourth-cluster showed association of genes *GMPS, ETFBKMT, PODXL, RAB34, PUS10, IL34, HDLBP, ITGA6, ZBTB7C, EIF3K, DBF4, NPHP3, STAT4, SRP72* and *SRBD1* in most cases downregulated in all ZIKV exposed infants (either CZS and ZIKV, without clinical signs) and upregulated in CTRL infants.

**Figure 5.**
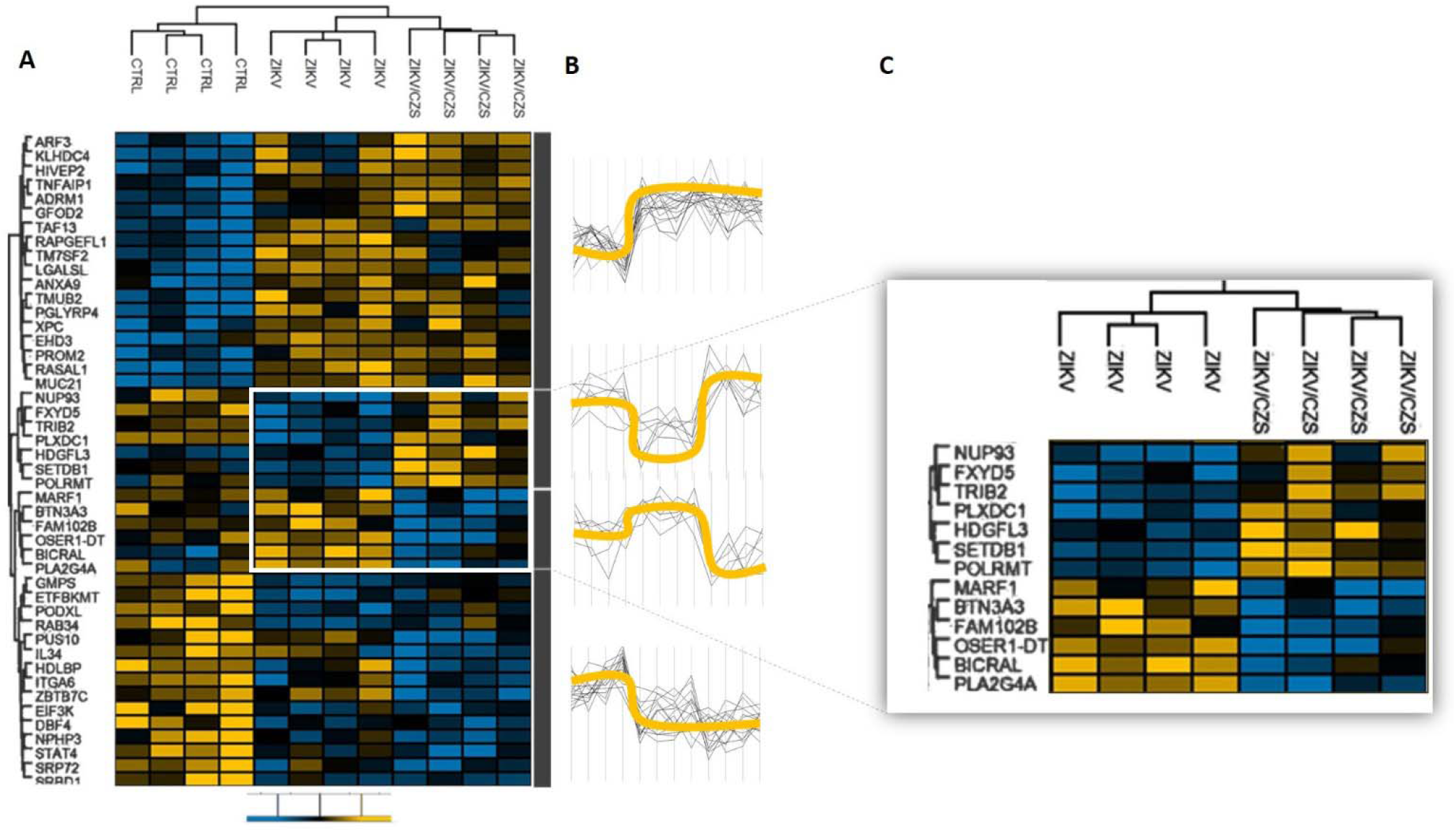
Heatmap representation of significant transcripts clusters (p < 0.001) in ZIKV/CZS, ZIKV without clinical signs and unaffected samples. **A**. Changes in expression levels are displayed from blue (downregulated) to yellow (upregulated). **B**. Expression levels between groups represented by yellow line. **C**. Highlighted are the differential gene expression between the ZIKV groups exposed. Heat map were generated, and hierarchical clustering was performed using Perseus based on log2 fold-change data. Scale bar = Z score.

### Functional enrichments

We found a significant enrichment of Gene Ontology (GO) annotations among the 63 differentially expressed transcripts detected in the RNA-Seq experiment (Figures 6 and 8). The top three most significantly enriched GO differentially regulated transcripts in ZIKV/CZS compared to CTRL (Supplemental Table 2) are “PI3K-Akt signaling pathway”, “Pathways in cancer”, “Mismatch repair” and “mTOR signaling pathway” (Figure 6). As the most significantly enriched GO from ZIKV compared to CTRL group are “DNA replication”, “Cell cycle” and “Non-homologous end-joining” (Figure 8; Supplemental Table 3).

**Figure 6.**
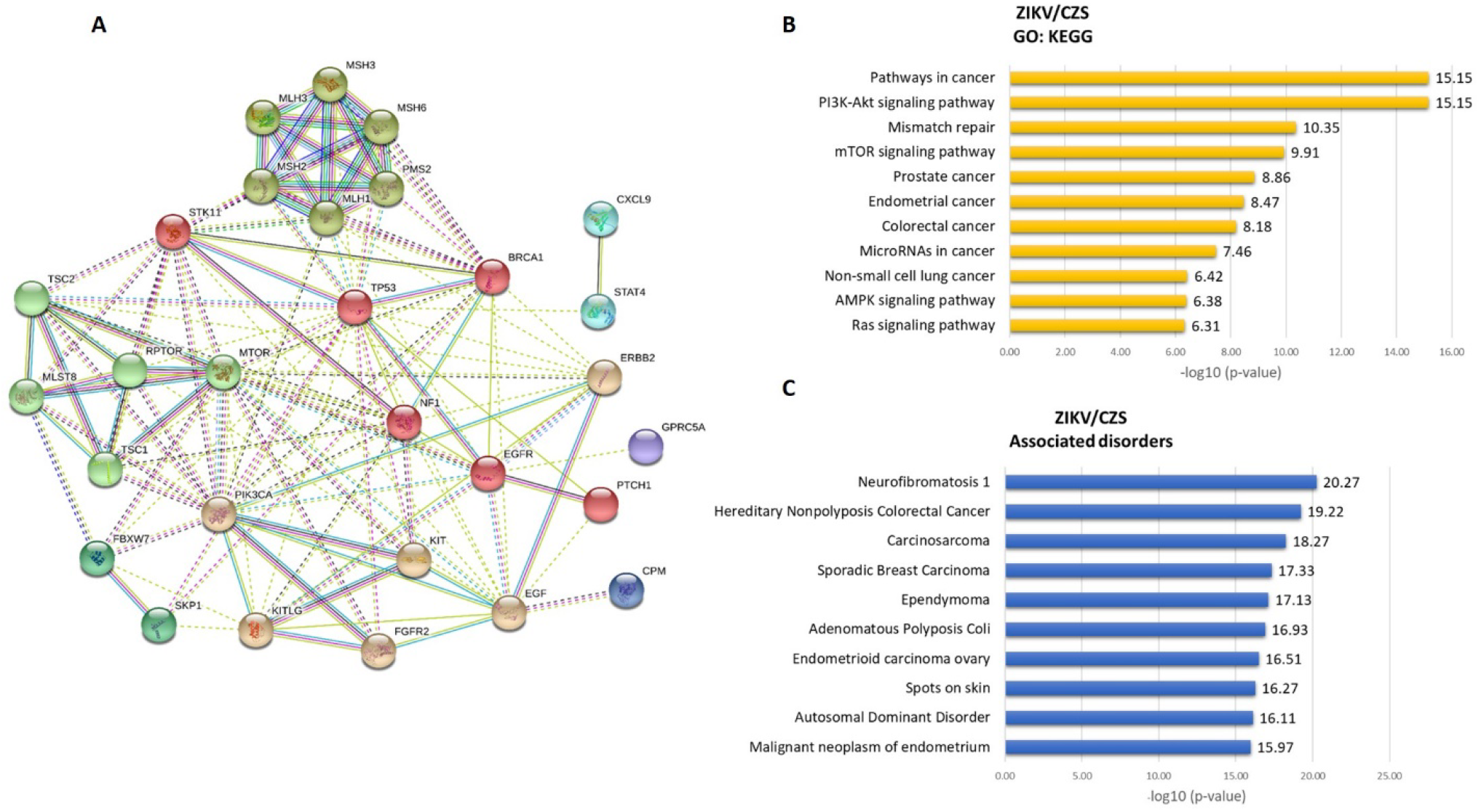
Deregulated genes in ZIKV/CZS babies. **A**. The thickness of the lines connecting the circles indicates the number of interactions between genes included in each Gene Ontology (GO) sub-category represented by each circle. **B**. Top 10 enriched GO interaction network according *STRING/KEGG* analysis. **C**. Top 10 associated disorders according *Toppgene* analysis.

**Figure 7.**
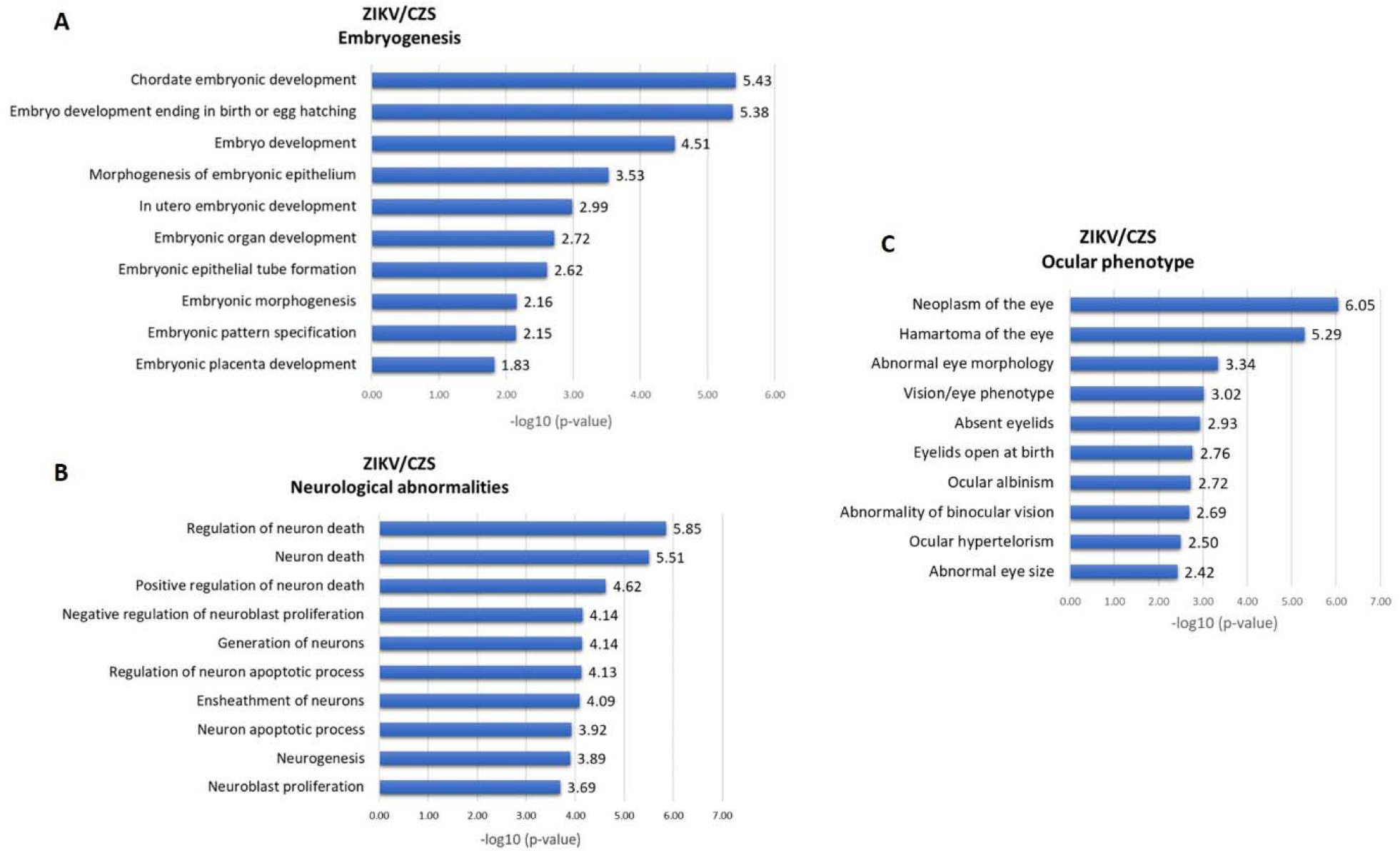
Congenital Zika Syndrome correlation gene expression / phenotype according *Toppgene* analysis. **A**. Top 10 embryogenesis stages related. Top 10 neurological process associated. C. Top 10 ocular phenotype associated.

**Figure 8.**
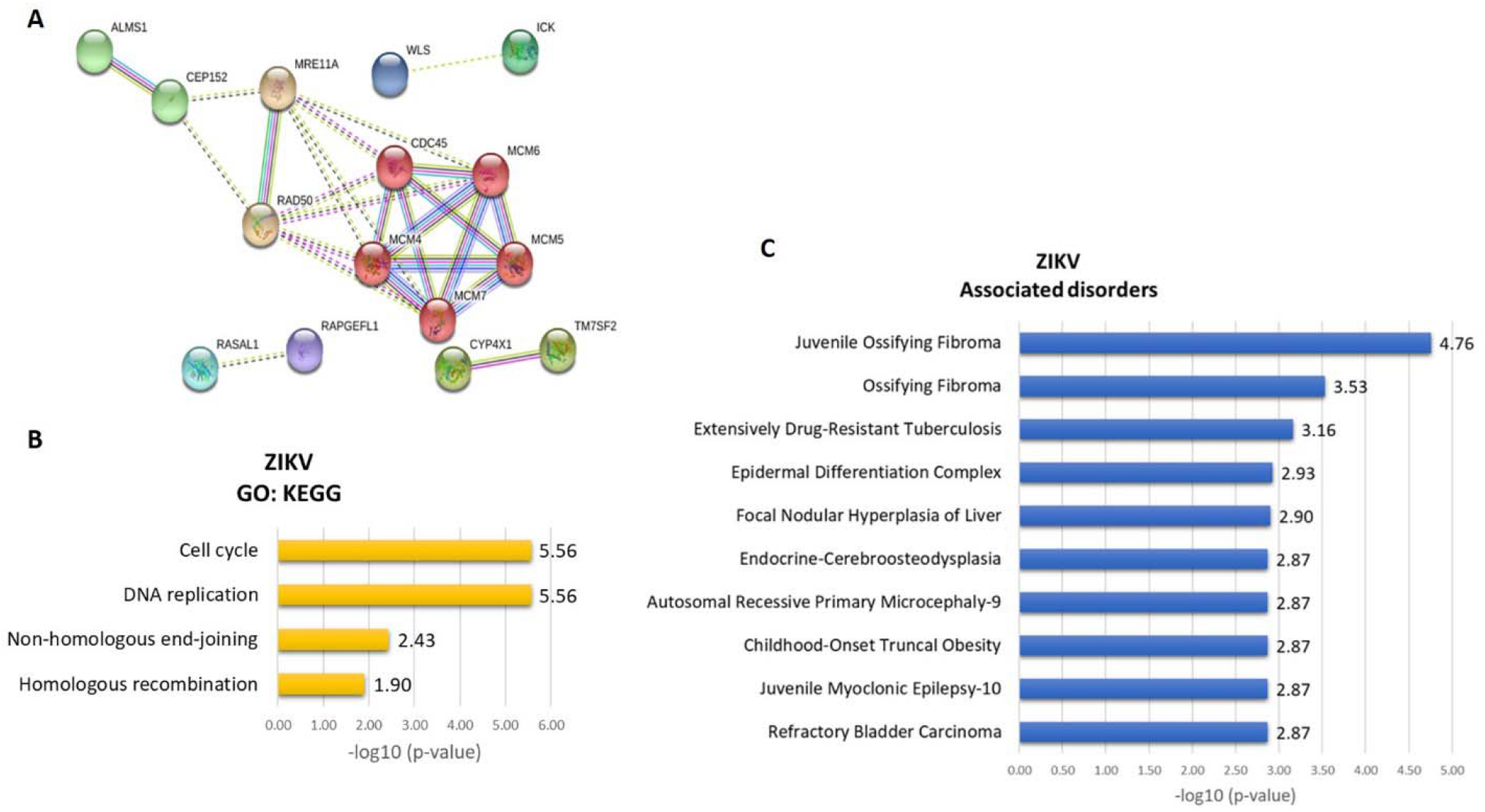
Asymptomatic ZiKV correlation gene expression / phenotype analysis. **A**. Deregulated genes in ZIKV babies. The thickness of the lines connecting the circles indicates the number of interactions between genes included in each Gene Ontology (GO) sub-category represented by each circle. **B**. Top 10 enriched GO interaction network according *STRING/KEGG* analysis. **C**. Top 10 associated disorders according *Toppgene* analysis.

## Discussion

### The eye as neuroepithelial model to study Zika virus infection

Human sensory organs and cranial sensory ganglia are functionally diverse but share a common developmental origin from neural progenitor cells. During the different stages of embryogenesis - through a process that requires a rigorous integration between multiple signaling pathways, repressive transcription factors, cell target specification and morphogenesis - the inner ear, olfactory epithelium, lens, and sensory neurons are gradually originated^32^. Besides, the eye – a specialized CNS compartment - has been proposed as a model for studies in neurodegenerative diseases^18,22.^ We have recently described a novel non-invasive method that enables collecting ocular cells from children^22^ as alternative neuroepithelial model to study Zika virus infection and here we applied this technique, for the first time, to investigate the extent of gene expression dysregulation in the ocular tissue of zika virus prenatally exposed children with and without CZS.

### Differentially expressed genes involved in neurogenesis, eye development, and neurological disorders

Molecular and cellular events leading to abnormal nervous system development in CZS infants are poorly understood, but the host’s response to ZIKV insults is likely to be modulated by the combined effects of multiple genes. Several genes emerged here as differentially expressed in both groups of exposed infants (CZS and ZIKV), many of which are involved in neurogenesis and ocular development (including mRNA expression in embryonic tissues and primordial stem cells). Indeed, the impact of ZIKV infection on neuronal regulation has already been shown experimentally^33,34,35, 36, 36,37^ and is consistent with the hypothesis that changes in central nervous system embryogenesis that predict adverse outcome as observed in CZS are modulated by deregulation in several signaling pathways (Figures 6 and 7; Supplemental Tables 4, 5 and 6). Here we describe several genes and their biological contexts associated to neurodevelopment (gene details^38^ in Boxes 1A, 1B and 2A) and highlight a likely implication of a deregulation of these molecules in CZS (Figures 6 and 7).

#### Box 1.

**1A.** Upregulated genes description. **1B.** Downregulated genes description.

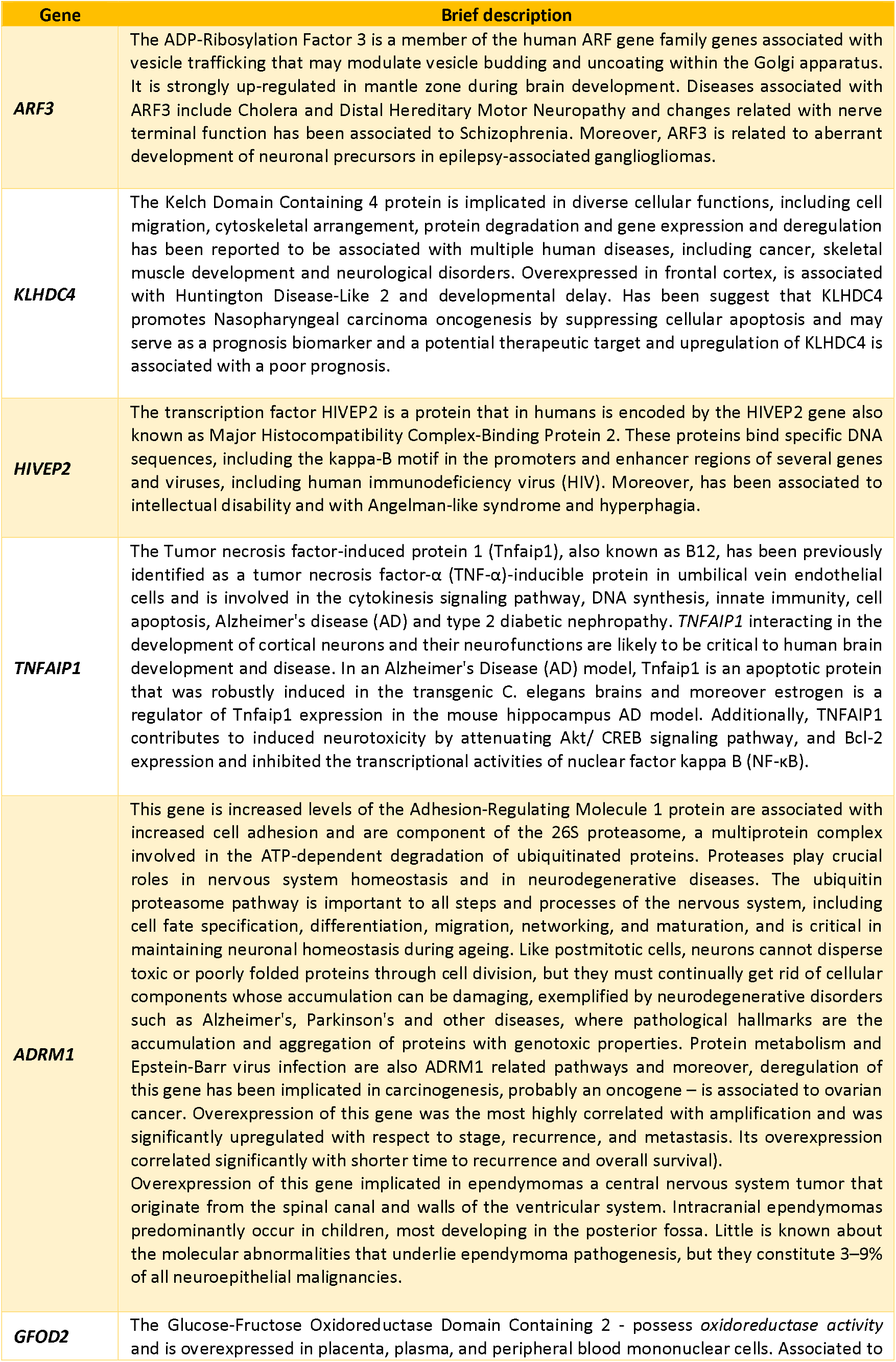
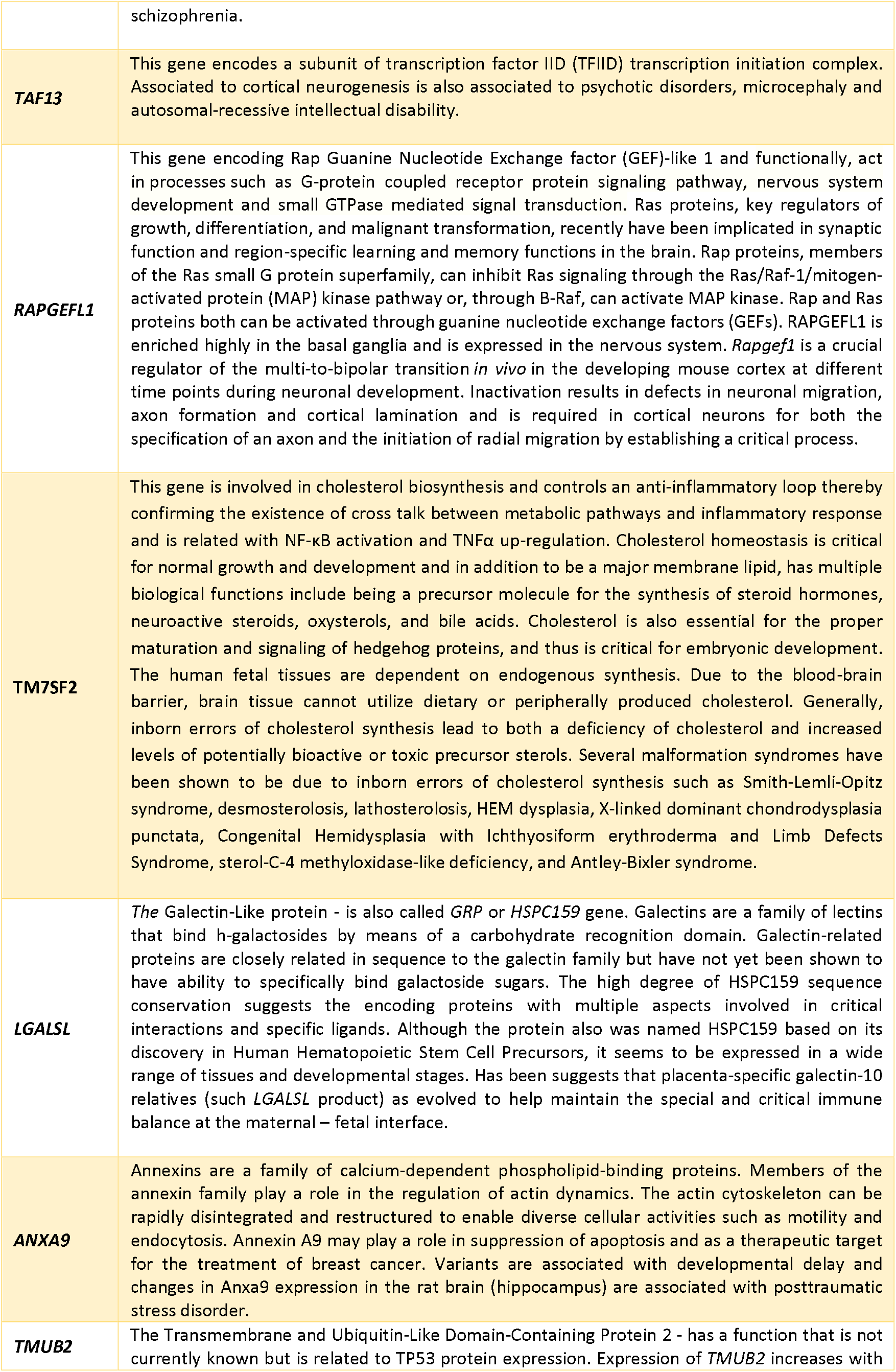
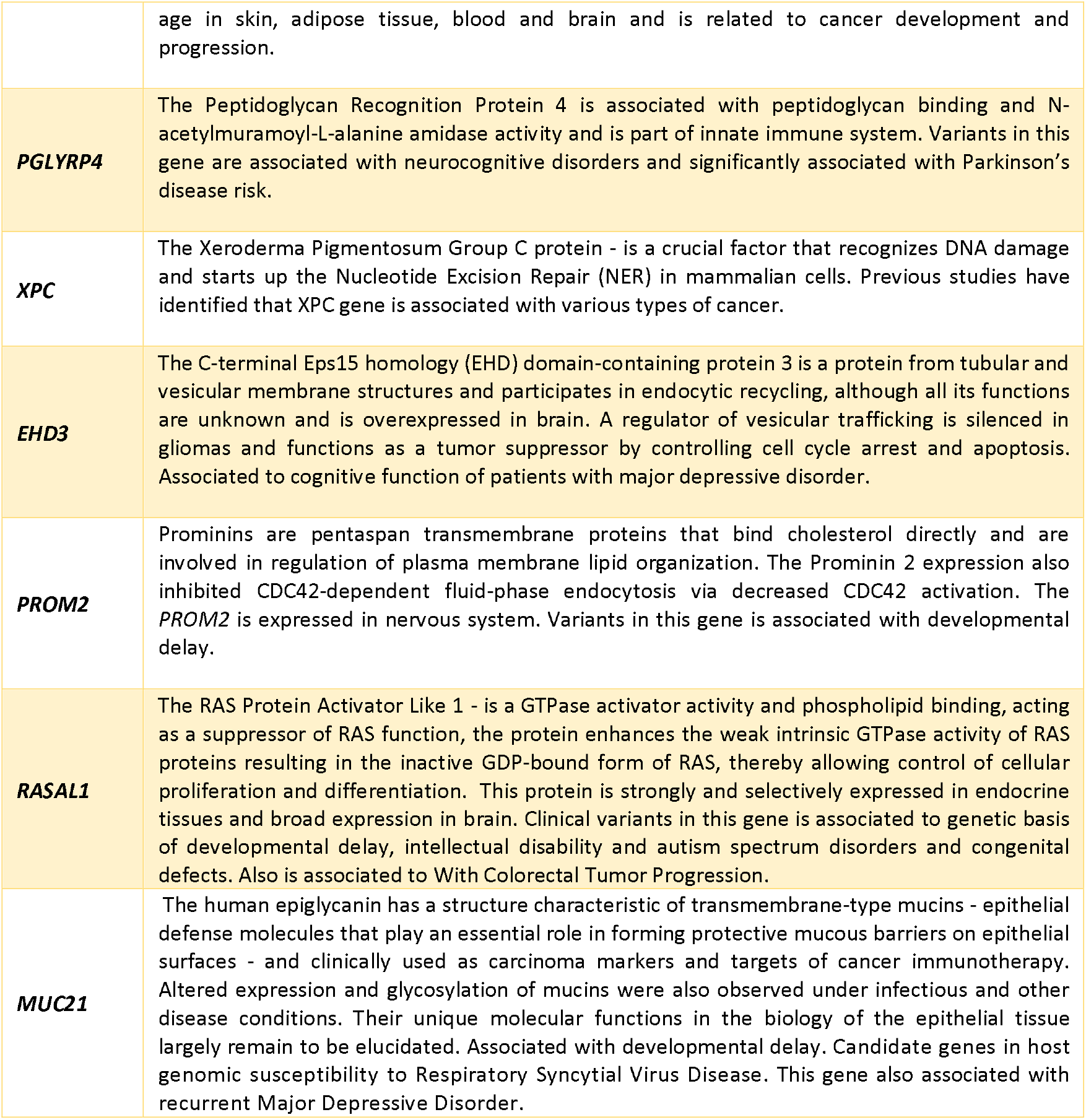

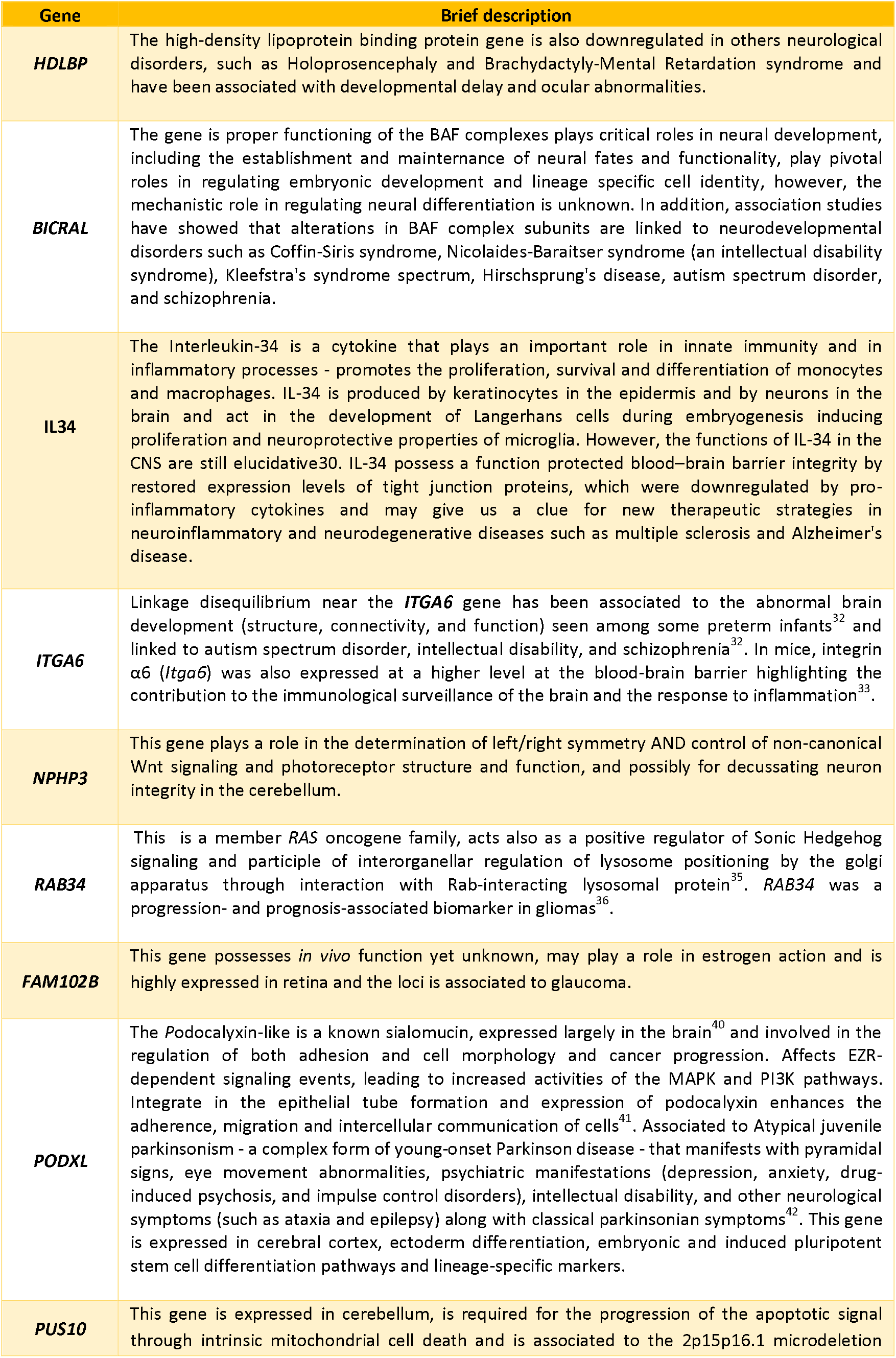
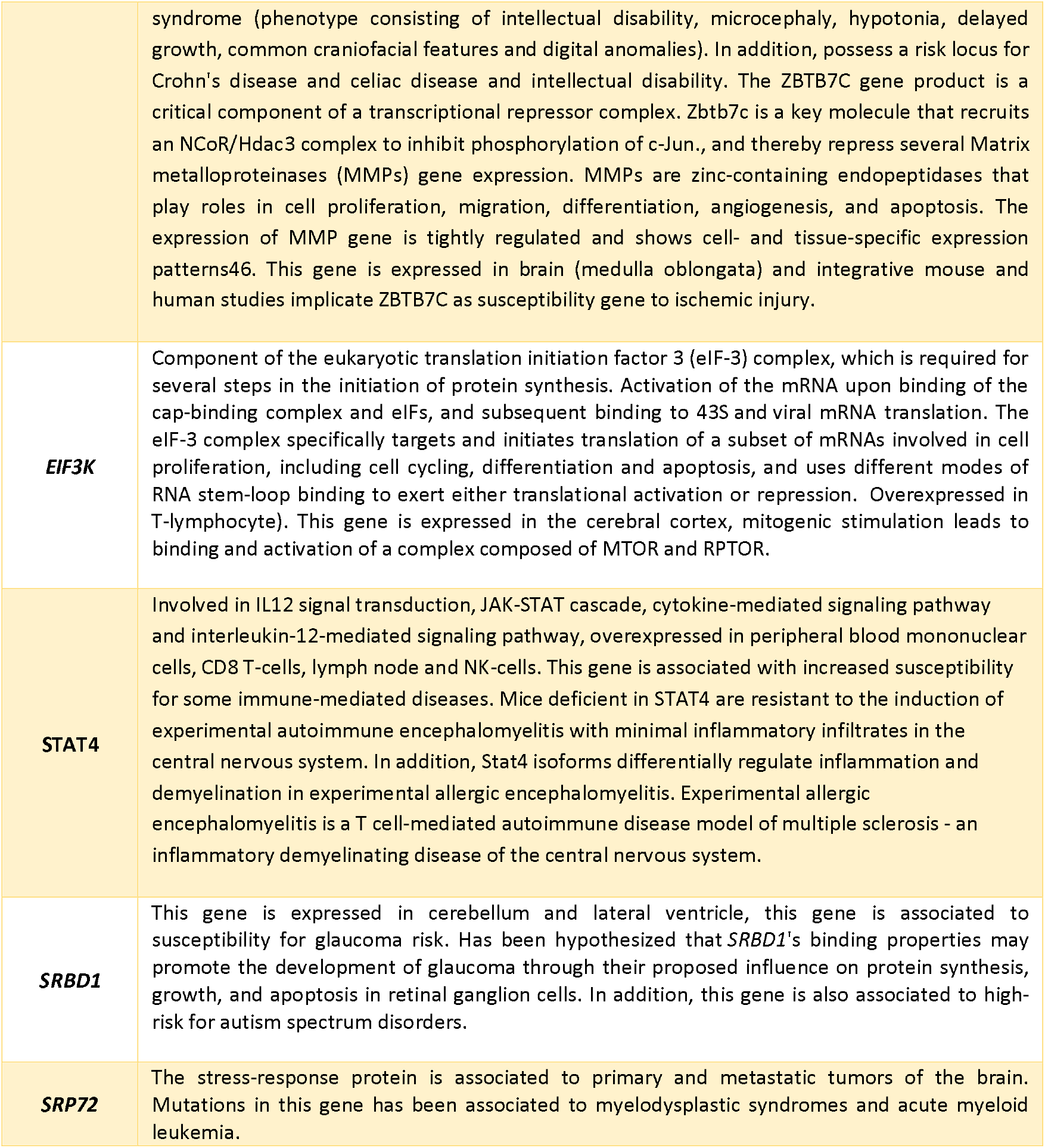

DOWNREGULATED GENES
| Gene | Brief description |
| --- | --- |
| ***HDLBP*** | The high-density lipoprotein binding protein gene is also downregulated in others neurological disorders, such as Holoprosencephaly and Brachydactyly-Mental Retardation syndrome and have been associated with developmental delay and ocular abnormalities. |
| ***BICRAL*** | The gene is proper functioning of the BAF complexes plays critical roles in neural development, including the establishment and maintenance of neural fates and functionality, play pivotal roles in regulating embryonic development and lineage specific cell identity, however, the mechanistic role in regulating neural differentiation is unknown. In addition, association studies have showed that alterations in BAF complex subunits are linked to neurodevelopmental disorders such as Coffin-Siris syndrome, Nicolaides-Baraitser syndrome (an intellectual disability syndrome), Kleefstra's syndrome spectrum, Hirschsprung's disease, autism spectrum disorder, and schizophrenia. |
| ***IL34*** | The Interleukin-34 is a cytokine that plays an important role in innate immunity and in inflammatory processes - promotes the proliferation, survival and differentiation of monocytes and macrophages. IL-34 is produced by keratinocytes in the epidermis and by neurons in the brain and act in the development of Langerhans cells during embryogenesis inducing proliferation and neuroprotective properties of microglia. However, the functions of IL-34 in the CNS are still elucidative<sup>30</sup>. IL-34 possess a function protected blood-brain barrier integrity by restored expression levels of tight junction proteins, which were downregulated by pro-inflammatory cytokines and may give us a clue for new therapeutic strategies in neuroinflammatory and neurodegenerative diseases such as multiple sclerosis and Alzheimer's disease. |
| ***ITGA6*** | Linkage disequilibrium near the ***ITGA6*** gene has been associated to the abnormal brain development (structure, connectivity, and function) seen among some preterm infants<sup>32</sup> and linked to autism spectrum disorder, intellectual disability, and schizophrenia<sup>32</sup>. In mice, integrin $\alpha 6$ (*Itga6*) was also expressed at a higher level at the blood-brain barrier highlighting the contribution to the immunological surveillance of the brain and the response to inflammation<sup>33</sup>. |
| ***NPHP3*** | This gene plays a role in the determination of left/right symmetry AND control of non-canonical Wnt signaling and photoreceptor structure and function, and possibly for decussating neuron integrity in the cerebellum. |
| ***RAB34*** | This is a member *RAS* oncogene family, acts also as a positive regulator of Sonic Hedgehog signaling and participate of interorganellar regulation of lysosome positioning by the golgi apparatus through interaction with Rab-interacting lysosomal protein<sup>35</sup>. *RAB34* was a progression- and prognosis-associated biomarker in gliomas<sup>36</sup>. |
| ***FAM102B*** | This gene possesses *in vivo* function yet unknown, may play a role in estrogen action and is highly expressed in retina and the loci is associated to glaucoma. |
| ***PODXL*** | The Podocalyxin-like is a known sialomucin, expressed largely in the brain<sup>40</sup> and involved in the regulation of both adhesion and cell morphology and cancer progression. Affects EZR-dependent signaling events, leading to increased activities of the MAPK and PI3K pathways. Integrate in the epithelial tube formation and expression of podocalyxin enhances the adherence, migration and intercellular communication of cells<sup>41</sup>. Associated to Atypical juvenile parkinsonism - a complex form of young-onset Parkinson disease - that manifests with pyramidal signs, eye movement abnormalities, psychiatric manifestations (depression, anxiety, drug-induced psychosis, and impulse control disorders), intellectual disability, and other neurological symptoms (such as ataxia and epilepsy) along with classical parkinsonian symptoms<sup>42</sup>. This gene is expressed in cerebral cortex, ectoderm differentiation, embryonic and induced pluripotent stem cell differentiation pathways and lineage-specific markers. |
| ***PUS10*** | This gene is expressed in cerebellum, is required for the progression of the apoptotic signal through intrinsic mitochondrial cell death and is associated to the 2p15p16.1 microdeletion |
| DOWNREGULATED GENES |  |
|  | syndrome (phenotype consisting of intellectual disability, microcephaly, hypotonia, delayed growth, common craniofacial features and digital anomalies). In addition, possess a risk locus for Crohn's disease and celiac disease and intellectual disability. The ZBTB7C gene product is a critical component of a transcriptional repressor complex. Zbtb7c is a key molecule that recruits an NCoR/Hdac3 complex to inhibit phosphorylation of c-Jun., and thereby repress several Matrix metalloproteinases (MMPs) gene expression. MMPs are zinc-containing endopeptidases that play roles in cell proliferation, migration, differentiation, angiogenesis, and apoptosis. The expression of MMP gene is tightly regulated and shows cell- and tissue-specific expression patterns<sup>46</sup>. This gene is expressed in brain (medulla oblongata) and integrative mouse and human studies implicate ZBTB7C as susceptibility gene to ischemic injury. |
| **EIF3K** | Component of the eukaryotic translation initiation factor 3 (eIF-3) complex, which is required for several steps in the initiation of protein synthesis. Activation of the mRNA upon binding of the cap-binding complex and eIFs, and subsequent binding to 43S and viral mRNA translation. The eIF-3 complex specifically targets and initiates translation of a subset of mRNAs involved in cell proliferation, including cell cycling, differentiation and apoptosis, and uses different modes of RNA stem-loop binding to exert either translational activation or repression. Overexpressed in T-lymphocyte). This gene is expressed in the cerebral cortex, mitogenic stimulation leads to binding and activation of a complex composed of MTOR and RPTOR. |
| **STAT4** | Involved in IL12 signal transduction, JAK-STAT cascade, cytokine-mediated signaling pathway and interleukin-12-mediated signaling pathway, overexpressed in peripheral blood mononuclear cells, CD8 T-cells, lymph node and NK-cells. This gene is associated with increased susceptibility for some immune-mediated diseases. Mice deficient in STAT4 are resistant to the induction of experimental autoimmune encephalomyelitis with minimal inflammatory infiltrates in the central nervous system. In addition, Stat4 isoforms differentially regulate inflammation and demyelination in experimental allergic encephalomyelitis. Experimental allergic encephalomyelitis is a T cell-mediated autoimmune disease model of multiple sclerosis - an inflammatory demyelinating disease of the central nervous system. |
| **SRBD1** | This gene is expressed in cerebellum and lateral ventricle, this gene is associated to susceptibility for glaucoma risk. Has been hypothesized that *SRBD1*'s binding properties may promote the development of glaucoma through their proposed influence on protein synthesis, growth, and apoptosis in retinal ganglion cells. In addition, this gene is also associated to high-risk for autism spectrum disorders. |
| **SRP72** | The stress-response protein is associated to primary and metastatic tumors of the brain. Mutations in this gene has been associated to myelodysplastic syndromes and acute myeloid leukemia. |

#### Box 2.

**2A.** CZS associated genes. **2B.** Asymptomatic ZIKV associated genes.

CONGENITAL ZIKA SYNDROME ASSOCIATED GENES
| Gene | Brief description |
| --- | --- |
| ***MARF1*** | Encodes a putative peroxisomal protein required to protect from DNA double-strand breaks, regulation of gene expression, negative regulation of phosphatase activity and cell differentiation. |
| ***BTN3A3*** | Butyrophilin is a group of major histocompatibility complex (MHC)-associated [PubMed 20208008]]. Plays a role in T-cell responses in the adaptive immune response, involved in cell proliferation and development, Immune system process, T cell mediated immunity, regulation of immune response, and T cell receptor signaling pathway. This gene is overexpressed in peripheral blood mononuclear cells, CD4 Tcells, CD8 Tcells, frontal cortex and spleen. |
| ***FAM102B*** | See details in Box 1 |
| ***OSER1-DT*** | The Oxidative Stress-Responsive 1 (Divergent Transcript) possess clinical variants associated developmental delay/intellectual disability, autism spectrum disorders or congenital anomalies. |
| ***BICRAL*** | See details in Box 1 |
| ***PLA2G4A*** | Encodes a member of the cytosolic phospholipase A2 group IV family. The enzyme catalyzes the hydrolysis of membrane phospholipids to release arachidonic acid which is subsequently metabolized into eicosanoids. Eicosanoids, including prostaglandins and leukotrienes, are lipid-based cellular hormones that regulate hemodynamics, inflammatory responses, and other intracellular pathways. Involved in G Protein-coupled Receptors Signaling, MAPK-Erk Pathway, P38 MAPK signaling pathway, signaling via JAK/STAT and MAPK cascades, Immune response CCR3 signaling in eosinophils, VEGF - VEGF R2 signaling pathways, all-trans-retinoic acid signaling in brain, and cAMP pathway. Transcripts in embryonic tissues and stem cells in cardiovascular system, blood brain barrier; gastrointestinal tract respiratory system, extraembryonic mesoderm and brain. |

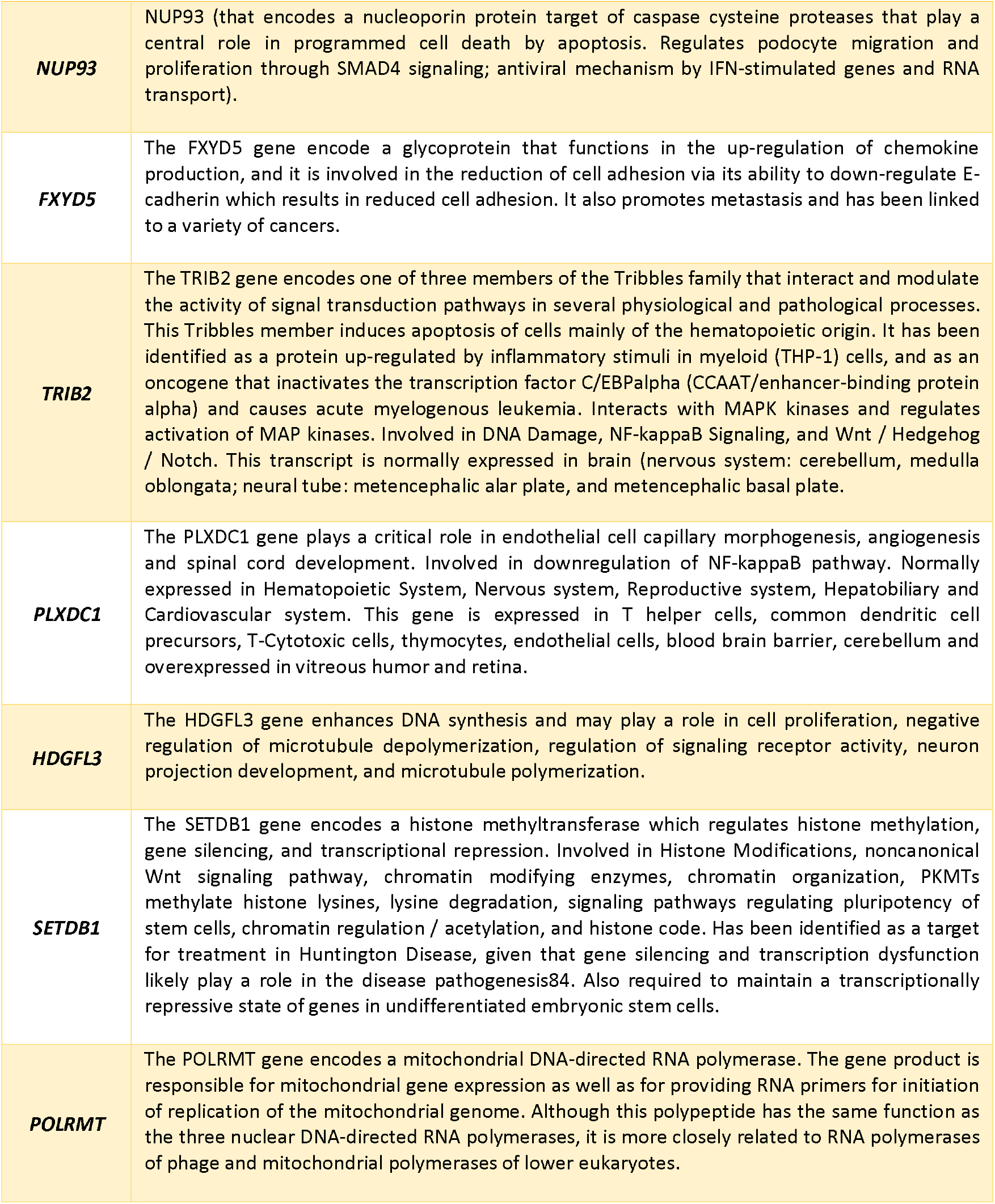

Collectively, the identity and function of these genes support the hypothesis that Zika virus infection in the first trimester of pregnancy affects precursor cells with effects that extend during the fetal developmental period and can still be seen in the differentiated stages of the cells in infants after birth (Figure 9). Ocular surface cells collected around 21 months after birth contain changes in embryonic marker genes and may be a representation of the interaction of precursor neuronal cells with the mechanisms of Zika virus infection during embryogenesis. Surprisingly, however, we also observed gene expression dysregulation of neurological genes in ZIKV groups associated to several neurological syndromes, developmental delay, intellectual disability, autism spectrum disorders, schizophrenia, and/or ocular impairment that we hypothesize might be relevant in these children. As demonstrated by several studies, there has been an advance in knowledge about the pathophysiology of ZIKV infection in the embryonic development of the CNS^33, 39, 34, 35, 36, 40, 37^. However, the knowledge about which pathways are associated with the pathogenic mechanism remains poorly understood. This directly interferes in the application of a more effective therapeutic approach, considering the serious clinical consequences for ZIKV exposed individuals^41^, and evidenced here for prenatally ZIKV exposed - both asymptomatic and CZS patients.

**Figure 9.**
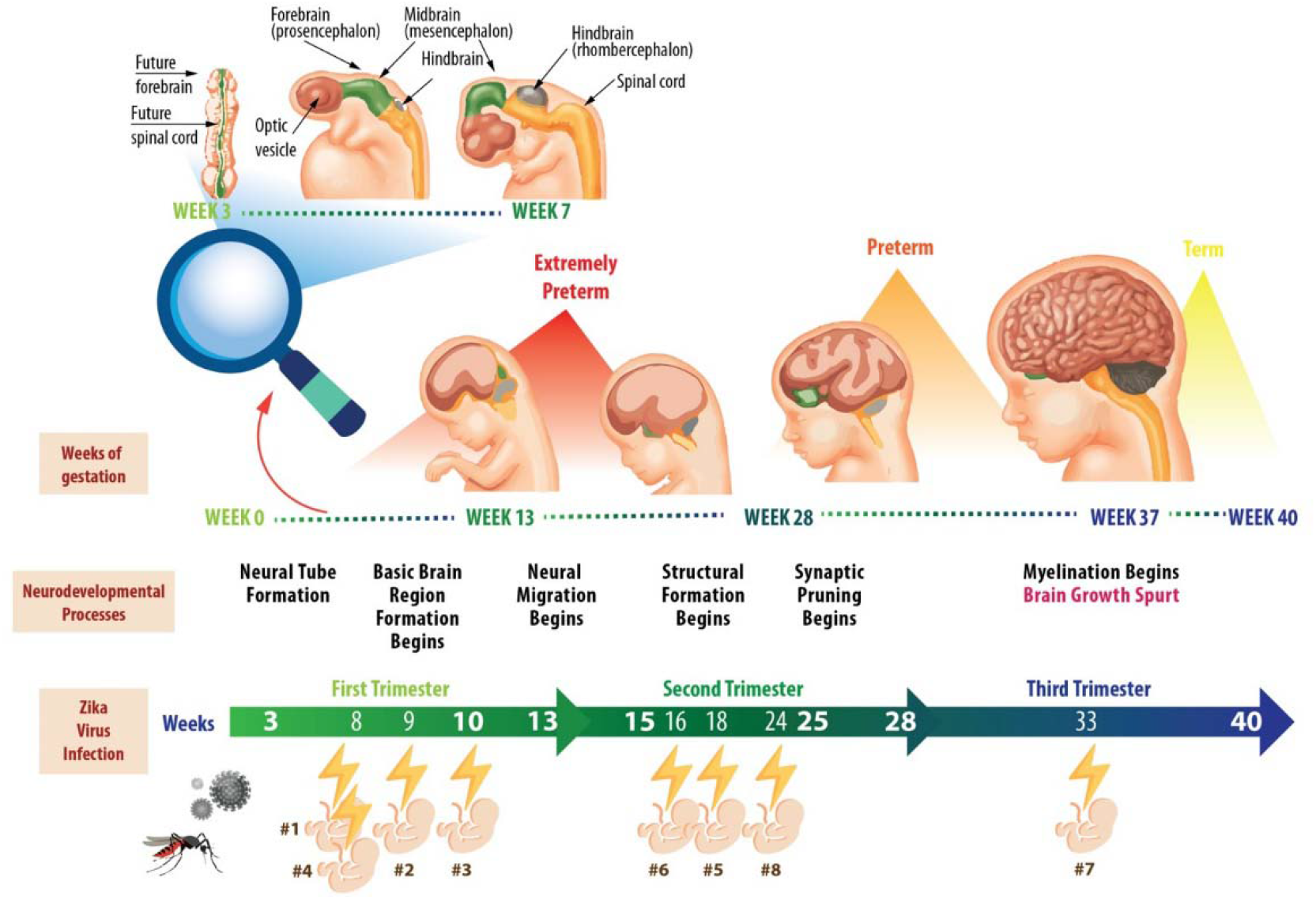
Schematic figure representing ZIKV prenatal exposure related to embryonic stages and developmental process in this study.

### Dysregulation of genes associated to Congenital Zika Syndrome

Comparative transcriptomics into ZIKV exposed infants and controls showed a downregulation of six genes also associated to CZS phenotypes (Figure 5). Here, we hypothesize that haploinsufficiency of these genes may have contributed to the CZS phenotype. Clinical variants of the ***OSER1-DT*** gene are associated with developmental delay/intellectual disability, autism spectrum disorders and congenital anomalies^42^; the ***BTN3A3*** gene is involved in immunity, cell proliferation and development^43^; the ***FAM102B*** gene possesses *in vivo* function unknown^38^ but is highly expressed in retina^44^ and associated to glaucoma^45^; the ***PLA2G4A*** gene is involved in inflammatory responses and all-trans-retinoic acid signaling in brain and moreover, transcripts are in embryonic tissues, blood brain barrier, extraembryonic mesoderm^46^; the ***MARF1*** gene required to protect from DNA double-strand breaks^38^ and the ***BICRAL*** gene, a component of chromatin remodeling^47^ (See more gene details^38^ in Box 2A).

### Genes related to response to infection mechanisms

We identified some deregulated genes in ZIKV exposed infants (either CZS or asymptomatic) associated to immune response mechanisms. In first cluster, for example, the upregulated *HIVEP2* gene possess enhancer elements of numerous viral promoters (such as those of SV40, CMV, or HIV1) and *ANXA9* gene - which expression is regulated by IL-22 in keratinocytes^48,49^. On the other hand, genes shown in fourth cluster are downregulated, such as *IL34* gene (related to innate immunity and in inflammatory processes^50^; *EIF3K* gene (overexpressed in T-lymphocyte) and *STAT4* gene (See more gene details^38^ in Boxes 1 and 2). Several studies showed to the association of these genes with virus infection, such as papilloma virus 16 (HPV16), HIV infection and life cycle, Maraba, Hepatitis C and West Nile virus^38^.

### PI3/AKT/mTOR signaling in CZS

We found upregulation of several members of the PI3/AKT/mTOR signaling pathway (*BRCA1, EGF, EGFR, FGFR2, KIT, KITLG, MLST8, MTOR, PIK3CA, RPTOR, STK11, TP53, TSC1* and *TSC2* genes) in ocular samples from CZS infants. PI3K-AKT signaling pathway has an important role in brain growth and it is the cause of many brain overgrowth disorders^51^. The PI3K / AKT pathway is crucial for normal neural stem cells in the brain, balancing the maintenance of their multipotent, renewing and proliferating rather than differentiating and becoming inactive^52,53,54^. Subsequent to an injury, neural stem cells enter a repair phase and express high PI3K levels to increase proliferation^55^. A critical protagonism for the CDK1-PDK1-PI3K/Akt kinase signaling pathway has been suggested in the regulation of self-renewal, differentiation, and somatic reprogramming^55,56^. As observed during mouse embryo development, AKT activity has a role in mitosis and myogenic and neural differentiation and/or survival^57^. The causes of microcephaly by ZIKV are not fully understood but recently, it is reported that ZIKV protein NS4A and NS4B inhibit Akt-mTOR signaling pathways, resulting in upregulation of autophagy, and promoting viral replication^58^. It is been showed that ZIKV infection of human fetal neural stem cells cause inhibition of the Akt-mTOR pathway, leading to defective neurogenesis and aberrant activation of autophagy^58^. ZIKV proteins to improve viral replication by blocking two innate pathways: interferon (IFN) and mTOR signaling (that normally would inhibit viral replication). The NS5 protein promotes degradation of the interferon effector STAT2, which would then promote transcription of interferon pathway genes. NS4A and NS4B inhibit mTOR signaling emanating from receptor tyrosine kinases. These effects of ZIKV proteins also to inhibit neurogenesis, endorsing cell death^59^.

### Dysregulation of DNA Repair and cellular death

We found upregulated Mismatch Repair (MMR) pathway (*MLH1, MLH3, MSH2, MSH3, MSH6* and *PMS2* genes) in ocular samples from CZS infants. It has been suggesting that DNA damage can cause premature differentiation and apoptosis^60^. For instance, the regulation of Msh2 protein levels in the ESC interferes the decision of engaging on DNA repair pathways or undergoing apoptosis^61^. High levels of Msh2 expression - as detected in CZS infants in our pilot study - may lead to apoptotic responses^62^. Here, we propose that genotoxic stress and DNA repair responses influence neurodegeneration in the CZS, considering the that poorly repaired DNA damage can be a trigger for apoptosis, and the consequent loss of neurons by apoptosis can result in neurodegeneration, and moreover persistent DNA damage can result in defective neurogenesis, such as observed in CZS. To date, no association has been demonstrated between MMR genes and Zika virus infection in patients. However, this relationship has already been shown experimentally^39^ and others studies on the role of MMR in viral infections. Levels of MSH2 and MLH1 mRNA were dramatically increased in cells after avian leukosis virus infection, suggest that viral infection may promote the increased expression of MSH2 and MLH1 genes^61^. Post-replicative MMR factors have been shown to be recruited into Epstein-Barr virus (EBV) replication compartments, repairing errors that arise during viral replication or inhibiting recombination between conflict sequences. EBV DNA polymerase has an intrinsic 3’ to 5’ exonuclease activity and the fidelity of viral genome replication is believed to be high, therefore the MMR system might increase the integrity of the viral genome rather than to activate cytotoxic responses^63^. For effective HSV-1 replication in normal human cells, MSH2 and MLH1 are equally required while MLH1 knockdown inhibits early viral gene expression and MSH2 seems to act at a later stage in the viral life cycle^64^. In fact, morphological changes associated with cell degeneration and death were observed in the cells of infants exposed to ZIKV, both in asymptomatic and in CZS, being predominant in this group. These results corroborate our molecular results and highlight the importance of further studies.

### Dysregulation in DNA replication, cell cycle and Non-homologous end-joining recombination

We identified downregulation of Minichromosome Maintenance protein complex (MCM) in asymptomatic ZIKV infants. MCM is a DNA helicase essential for genomic DNA replication that consists of six gene products, MCM2–7, which form a heterohexamer^65^. As a critical protein for cell division, MCM is also the target of various checkpoint pathways and deregulation of MCM function has been linked to genomic instability and a variety of carcinomas^65^. In zebrafish experimental model has demonstrated that mcm5 is down regulated in differentiated cells but is preserved in retinal stem cells regions. A regular depletion of maternally derived protein leads to an extended S phase, failure to exit cell cycle, apoptosis, and cell number decrease in *mcm5* mutant embryos. However, apoptosis increases only in the retina, tectum, and hindbrain but not in other late-proliferating tissues - around the third day of development - suggesting that different tissues may employ distinct cellular programs in response to MCM5 depletion^66^. We also identified a downregulation *of CDC45* a member of the highly conserved multiprotein complex which interacts with MCM7 and appears involved in mortality/aging, cellular phenotype, and embryo phenotype. The mRNA expression is detected in embryonic tissues and stem cells from neural tube, mesencephalic ventricular zone, telencephalon, diencephalic ventricular zone, metencephalic alar plate and brain, choroid plexus and medulla oblongata and phenotypes associated included cognitive impairment and cognitive decline measurement^38^. Additionally, we also detected a downregulation of *MRE11A* gene that encodes a nuclear protein involved in homologous recombination, telomere length maintenance, and DNA double-strand break repair. As part of the MRN complex (a complex of three proteins — **M**re11, **R**ad50 and **N**bs1), it interacts with MCM9 and the interaction recruits the complex to DNA repair sites^67^. A component of the BASC complex (BRCA1-associated genome surveillance complex), at least composed of BRCA1, MSH2, MSH6, MLH1, ATM, BLM, RAD50, MRE11 and NBN^68^, interacts with herpes simplex virus 1 protein UL12^69^, and in E4 adenovirus infection is inactivated and degraded by viral oncoproteins, thereby preventing concatenation of viral genomes in infected cells. The MRE11A gene also is associated to increased risk of cancer^70^,^71^.

### Relevance for clinical surveillance

In this pilot study we highlight the cellular and molecular changes and the potential clinical implications in infants exposed prenatally to the Zika virus, not only in CZS, but also for those without clinical signs - asymptomatic (Figure 8; supplemental Table 7). Indeed, the results found alert for surveillance after a viral exposure, especially in asymptomatic individuals, considering the potential risk of a new Zika virus epidemic^3^. We identified deregulated genes and pathways involved in developmental disorders and cancer risk in CZS infants (Figure 6; Supplemental Table 8). Several parallelisms exist between development and cancer due to the similarities between normal tissue stem cells and cancer stem/progenitor cells^72^. Several developmental disorders are associated with congenital malformations and cancer risk and the risk is higher in children with congenital anomalies and some specific genetic syndromes^73^. Nevertheless, little is known regarding how defects in these genes and signaling pathways can produce congenital developmental abnormalities as well as alterations underlying the development of neoplastic process. Tumorigenesis associated with developmental syndromes is related to several variables such as pathways, genes, and genetic/epigenetic alterations as well as the origin of these mutations (somatic or germinative). However, despite the small number of samples evaluated in this pilot study, the monitoring and screening for pediatric cancer predisposition syndromes^74^ should not be discarded in CZS infants.

## Conclusion

Ocular cells used for the first time as a neuroepithelial model revealed the effect of Zika infection on primordial neuronal cell genes, evidenced by changes in genes associated with embryonic cells. Precisely because this is a pilot study, further studies with a larger sampling should be performed to corroborate these findings. Despite the small number of samples evaluated, the results described here support those previously described in the literature about Zika-host interaction. The changes in the genes validate an association with the gestational period of the occurrence of the infection and testify to the resulting clinical and ophthalmological pathologies. On the other hand, evidence that there is a deregulation in genes related to cell death brings to the fore the concern with the early onset of other associated pathologies. Evidence of deregulated genes associated with cancer raises concerns about the surveillance regarding the appearance of tumors. Here we support the hypothesis that the CNS changes in infants with CZS may be occurring beyond the consequences of the congenital infectious process. Thus, it will be interesting to study whether regulatory changes of the primordial genes of development during embryogenesis, as detected in this pilot study, might contribute to the neuropathological condition observed in this syndrome.

## Data Availability

The data for this study have been deposited in the European Nucleotide Archive at EMBL EBI under accession number PRJEB40869. Additional data supporting the findings of this study are available within the article and its Supplemental files or are available from the authors upon request. Due to privacy and consent restrictions, individualized RNAseq data files cannot be uploaded publicly, but might be available to researchers upon request and signature of a Data Use Agreement.

https://www.ebi.ac.uk/ena/browser/view/PRJEB40869

## Data availability

The data for this study have been deposited in the European Nucleotide Archive (ENA) at EMBL-EBI under accession number PRJEB40869 (https://www.ebi.ac.uk/ena/browser/view/PRJEB40869). Additional data supporting the findings of this study are available within the article and its Supplemental files or are available from the authors upon request. Due to privacy and consent restrictions, individualized RNAseq data files cannot be uploaded publicly, but might be available to researchers upon request and signature of a Data Use Agreement.

## Acknowledgements

We are grateful to Prof. Lemos for helping us with the initial steps in the RNAseq analysis tools and for reviewing of the first versions the manuscript. We thank the Zika Study Group of the Clinical Research Unit from the Universidade Federal Fluminense especially to the patients and their families. We dedicate this work to the Super Special Mom Mrs. Simone Duarte and to our sweet Guilherme (*in memoriam*).

## Author contributions

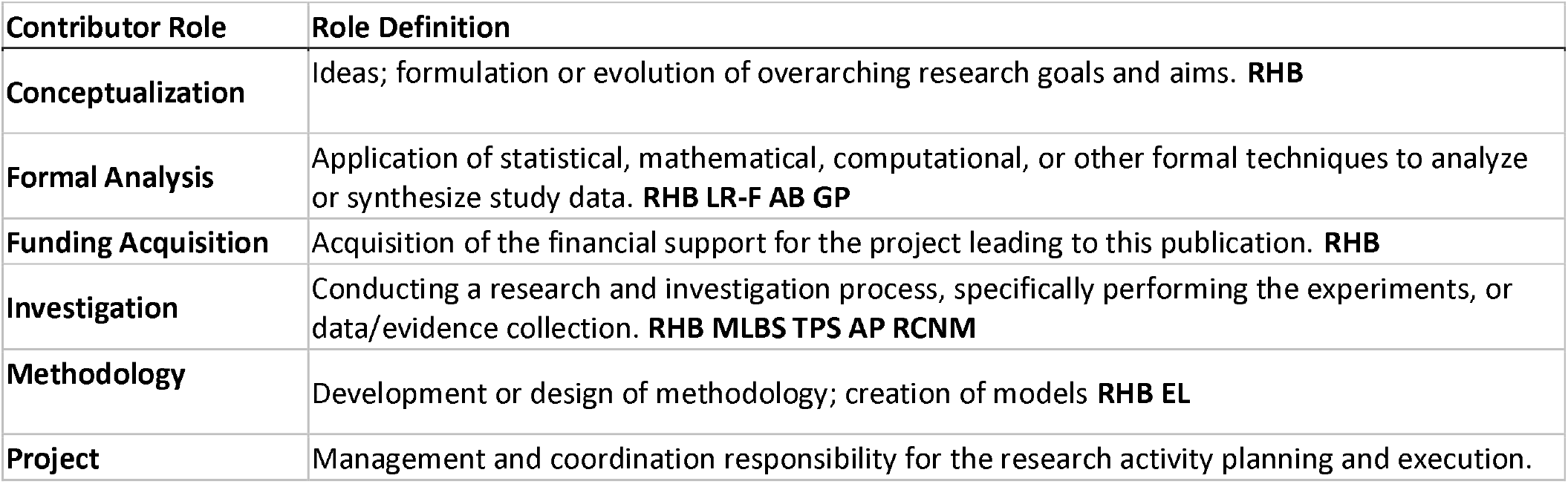
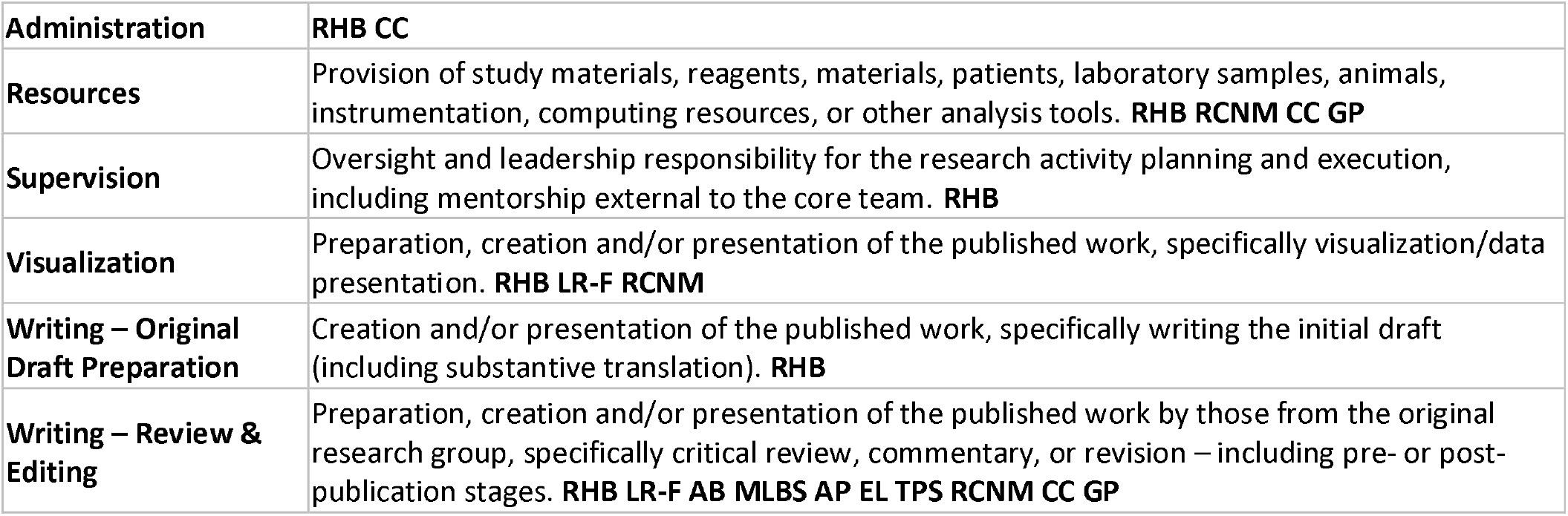

## Tables

**Table 1. Descriptive table of expression differentiated of transcripts into ZIKV/CZS and CTRL babies**.

**Table 2. Descriptive table of expression differentiated of transcripts into asymptomatic ZIKV and CTRL babies**.

**Supplemental Table 1. Clinical data**.

**Supplemental Table 2. Deregulated genes in ZIKV/CZS babies, enriched GO interaction network according *STRING/KEGG* analysis**.

**Supplemental Table 3. Deregulated genes in Asymptomatic ZIKV babies, enriched GO interaction network according *STRING/KEGG* analysis**.

**Supplemental Table 4. Correlation gene expression / Congenital Zika Syndrome phenotype according embryogenesis stages related. Results from *Toppgene* analysis**.

**Supplemental Table 5. Correlation gene expression / Congenital Zika Syndrome phenotype according neurological process associated. Results from *Toppgene* analysis**.

**Supplemental Table 6. Correlation gene expression / Congenital Zika Syndrome phenotype according ocular phenotype associated. Results from *Toppgene* analysis**.

**Supplemental Table 7. Correlation gene expression / Asymptomatic ZIKV phenotype according associated disorders. Results from *Toppgene* analysis**.

**Supplemental Table 8. Correlation gene expression / Congenital Zika Syndrome phenotype according associated disorders. Results from *Toppgene* analysis**.

## References

1. Kass DE, Merlino M. Zika Virus. N Engl J Med. 2016;375(3):294. https://www.who.int/newsroom/fact-sheets/detail/zika-virus. accessed July 18, 2020.

2. Campos GS, Bandeira AC, Sardi SI. Zika virus outbreak, Bahia, Brazil. Emerg Infect Dis. 2015;21(10):1885–1886. doi:10.3201/eid2110.150847

3. Kasprzykowski JI, Fukutani KF, Fabio H, et al. A recursive sub-typing screening surveillance system detects the appearance of the ZIKV African lineage in Brazil: Is there a risk of a new epidemic? Int J Infect Dis. 2020;96:579–581. doi:10.1016/j.ijid.2020.05.090

4. Mlakar J, Korva M, Tul N, et al. Zika Virus Associated with Microcephaly. N Engl J Med. 2016;374(10):951–958. doi:10.1056/NEJMoa1600651

5. Krauer F, Riesen M, Reveiz L, et al. Zika Virus Infection as a Cause of Congenital Brain Abnormalities and Guillain–Barré Syndrome: Systematic Review. PLoS Med. 2017;14(1). doi:10.1371/journal.pmed.1002203

6. Driggers RW, Ho C-Y, Korhonen EM, et al. Zika Virus Infection with Prolonged Maternal Viremia and Fetal Brain Abnormalities. N Engl J Med. 2016;374(22):2142-2151. doi:10.1056/NEJMoa1601824

7. Oliveira Melo AS, Malinger G, Ximenes R, Szejnfeld PO, Alves Sampaio S, Bispo De Filippis AM. Zika virus intrauterine infection causes fetal brain abnormality and microcephaly: Tip of the iceberg? Ultrasound Obstet Gynecol. 2016;47(1):6–7. doi:10.1002/uog.15831

8. Ventura C V., Maia M, Bravo-Filho V, Góis AL, Belfort R. Zika virus in Brazil and macular atrophy in a child with microcephaly. Lancet. 2016;387(10015):228. doi:10.1016/S0140-6736(16)00006-4

9. Ventura C V., Maia M, Ventura B V., et al. Ophthalmological findings in infants with microcephaly and presumable intra-uterus Zika virus infection. Arq Bras Oftalmol. 2016;79(1):1–3. doi:10.5935/0004-2749.20160002

10. Zin AA, Tsui I, Rossetto J, et al. Screening criteria for ophthalmic manifestations of congenital zika virus infection. JAMA Pediatr. 2017;171(9):847–854. doi:10.1001/jamapediatrics.2017.1474

11. De Paula Freitas B, De Oliveira Dias JR, Prazeres J, et al. Ocular findings in infants with microcephaly associated with presumed zika virus congenital infection in Salvador, Brazil. JAMA Ophthalmol. 2016;134(5):529–535. doi:10.1001/jamaophthalmol.2016.0267

12. Ventura C V., Ventura LO. Ophthalmologic manifestations associated with Zika Virus infection. Pediatrics. 2018;141(Suppl 2):S161–S166. doi:10.1542/peds.2017-2038E

13. Ventura C V., Maia M, Dias N, Ventura LO, Belfort R. Zika: Neurological and ocular findings in infant without microcephaly. Lancet. 2016;387(10037):2502. doi:10.1016/S0140-6736(16)30776-0

14. Fernandez MP, Parra Saad E, Ospina Martinez M, et al. Ocular histopathologic features of congenital Zika syndrome. JAMA Ophthalmol. 2017;135(11):1163–1169. doi:10.1001/jamaophthalmol.2017.3595

15. Miner JJ, Sene A, Richner JM, et al. Zika Virus Infection in Mice Causes Panuveitis with Shedding of Virus in Tears. Cell Rep. 2016;16(12):3208–3218. doi:10.1016/j.celrep.2016.08.079

16. Swaminathan S, Schlaberg R, Lewis J, Hanson KE, Couturier MR. Fatal Zika Virus Infection with Secondary Nonsexual Transmission. N Engl J Med. 2016;375(19):1907–1909. doi:10.1056/NEJMc1610613

17. Tan JJL, Balne PK, Leo YS, Tong L, Ng LFP, Agrawal R. Persistence of Zika virus in conjunctival fluid of convalescence patients. Sci Rep. 2017;7(1):1–5. doi:10.1038/s41598-017-09479-5

18. London A, Benhar I, Schwartz M. The retina as a window to the brain - From eye research to CNS disorders. Nat Rev Neurol. 2013;9(1):44–53. doi:10.1038/nrneurol.2012.227

19. Bandyopadhyay D, Qureshi A, Ashish K, Hajra A, Chakraborty S. Effect of ZIKA virus on adult eyes. Eur J Intern Med. 2018;47:e18–e19. doi:10.1016/j.ejim.2017.06.016

20. Lebov JF, Brown LM, MacDonald PDM, et al. Review: Evidence of Neurological Sequelae in Children With Acquired Zika Virus Infection. Pediatr Neurol. 2018;85:16–20. doi:10.1016/j.pediatrneurol.2018.03.001

21. L C, S MP, E A-P, et al. Persistence of Zika Virus After Birth: Clinical, Virological, Neuroimaging, and Neuropathological Documentation in a 5-Month Infant With Congenital Zika Syndrome. J Neuropathol Exp Neurol. 2018;77(3). doi:10.1093/JNEN/NLX116

22. Barbosa RH, dos Santos MLB, Silva TP, et al. Impression Cytology Is a Non-invasive and Effective Method for Ocular Cell Retrieval of Zika Infected Babies: Perspectives in OMIC Studies. Front Mol Neurosci. 2019;12:279. doi:10.3389/fnmol.2019.00279

23. STAR: ultrafast universal RNA-seq aligner | Bioinformatics | Oxford Academic. https://academic.oup.com/bioinformatics/article/29/1/15/272537. Accessed July 4, 2020.

24. Trapnell C, Roberts A, Goff L, et al. Differential gene and transcript expression analysis of RNA-seq experiments with TopHat and Cufflinks. Nat Protoc. 2012;7(3):562–578. doi:10.1038/nprot.2012.016

25. Anders S, Huber W. Differential expression analysis for sequence count data. Genome Biol. 2010;11(10):R106. doi:10.1186/gb-2010-11-10-r106

26. Love MI, Huber W, Anders S. Moderated estimation of fold change and dispersion for RNA-seq data with DESeq2. Genome Biol. 2014;15(12):550. doi:10.1186/s13059-014-0550-8

27. Eden E, Navon R, Steinfeld I, Lipson D, Yakhini Z. GOrilla: A tool for discovery and visualization of enriched GO terms in ranked gene lists. BMC Bioinformatics. 2009;10(1):48. doi:10.1186/1471-2105-10-48

28. Perseus. https://maxquant.net/perseus/. Accessed July 4, 2020.

29. STRING: functional protein association networks. https://string-db.org/. Accessed July 4, 2020.

30. Welcome to ToppGene. https://toppgene.cchmc.org/. Accessed July 4, 2020.

31. Rosa-Fernandes L, Hora Barbosa R, Luiza dos Santos MB, et al. CImP: Cellular Imprinting Proteomics applied to ocular disorders elicited by Congenital Zika virus Syndrome. bioRxiv. May 2019:648600. doi:10.1101/648600

32. Lleras-Forero L, Streit A. Development of the sensory nervous system in the vertebrate head: The importance of being on time. Curr Opin Genet Dev. 2012;22(4):315–322. doi:10.1016/j.gde.2012.05.003

33. Rosa-Fernandes L, Cugola FR, Russo FB, et al. Zika virus impairs neurogenesis and synaptogenesis pathways in human neural stem cells and neurons. Front Cell Neurosci. 2019;13. doi:10.3389/fncel.2019.00064

34. Hughes BW, Addanki KC, Sriskanda AN, McLean E, Bagasra O. Infectivity of Immature Neurons to Zika Virus: A Link to Congenital Zika Syndrome. EBioMedicine. 2016;10:65–70. doi:10.1016/j.ebiom.2016.06.026

35. Russo FB, Jungmann P, Beltrão-Braga PCB. Zika infection and the development of neurological defects. Cell Microbiol. 2017;19(6). doi:10.1111/cmi.12744

36. Goodfellow FT, Willard KA, Stice SL, Brindley MA, Wu X, Scoville S. Strain-dependent consequences of zika virus infection and differential impact on neural development. Viruses. 2018;10(10). doi:10.3390/v10100550

37. Ledur PF, Karmirian K, Pedrosa C da SG, et al. Zika virus infection leads to mitochondrial failure, oxidative stress and DNA damage in human iPSC-derived astrocytes. Sci Rep. 2020;10(1). doi:10.1038/s41598-020-57914-x

38. GeneCards - Human Genes | Gene Database | Gene Search. https://www.genecards.org/. Accessed July 4, 2020.

39. Park T, Kang MG, Baek SH, Lee CH, Park D. Zika virus infection differentially affects genome-wide transcription in neuronal cells and myeloid dendritic cells. PLoS One. 2020;15(4). doi:10.1371/journal.pone.0231049

40. Shao Q, Herrlinger S, Yang SL, et al. Zika virus infection disrupts neurovascular development and results in postnatal microcephaly with brain damage. Dev. 2016;143(22):4127–4136. doi:10.1242/dev.143768

41. Gratton R, Tricarico PM, Agrelli A, et al. In vitro zika virus infection of human neural progenitor cells: Meta-analysis of RNA-seq assays. Microorganisms. 2020;8(2). doi:10.3390/microorganisms8020270

42. Miller DT, Adam MP, Aradhya S, et al. Consensus Statement: Chromosomal Microarray Is a First-Tier Clinical Diagnostic Test for Individuals with Developmental Disabilities or Congenital Anomalies. Am J Hum Genet. 2010;86(5):749–764. doi:10.1016/j.ajhg.2010.04.006

43. Smith IA, Knezevic BR, Ammann JU, et al. BTN1A1, the Mammary Gland Butyrophilin, and BTN2A2 Are Both Inhibitors of T Cell Activation. J Immunol. 2010;184(7):3514–3525. doi:10.4049/jimmunol.0900416

44. harmonizome: a collection of processed datasets gathered to serve and mine knowledge about genes and proteins | Database | Oxford Academic. https://academic.oup.com/database/article/doi/10.1093/database/baw100/2630482. Accessed July 4, 2020.

45. Genetics of glaucoma | Human Molecular Genetics | Oxford Academic. https://academic.oup.com/hmg/article/26/R1/R21/3827806. Accessed July 4, 2020.

46. PLA2G4A gene - Genetics Home Reference - NIH. https://ghr.nlm.nih.gov/gene/PLA2G4A. Accessed July 4, 2020.

47. Sokpor G, Xie Y, Rosenbusch J, Tuoc T. Chromatin remodeling BAF (SWI/SNF) complexes in neural development and disorders. Front Mol Neurosci. 2017;10:243. doi:10.3389/fnmol.2017.00243

48. Wolk K, Witte E, Wallace E, et al. IL-22 regulates the expression of genes responsible for antimicrobial defense, cellular differentiation, and mobility in keratinocytes: A potential role in psoriasis. Eur J Immunol. 2006;36(5):1309–1323. doi:10.1002/eji.200535503

49. Wolk K, Witte E, Wallace E, et al. IL-22 regulates the expression of genes responsible for antimicrobial defense, cellular differentiation, and mobility in keratinocytes: A potential role in psoriasis. Eur J Immunol. 2006;36(5):1309–1323. doi:10.1002/eji.200535503

50. Ge Y, Huang M, Yao YM. Immunomodulation of interleukin-34 and its potential significance as a disease biomarker and therapeutic target. Int J Biol Sci. 2019;15(9):1835–1845. doi:10.7150/ijbs.35070

51. Hevner RF. Brain overgrowth in disorders of RTK-PI3K-AKT signaling: A mosaic of malformations. Semin Perinatol. 2015;39(1):36–43. doi:10.1053/j.semperi.2014.10.006

52. Wang L, Zhou K, Fu Z, et al. Brain Development and Akt Signaling: the Crossroads of Signaling Pathway and Neurodevelopmental Diseases. J Mol Neurosci. 2017;61(3):379–384. doi:10.1007/s12031-016-0872-y

53. Lee DY. Roles of mTOR Signaling in Brain Development. Exp Neurobiol. 2015;24(3):177–185. doi:10.5607/en.2015.24.3.177

54. Rafalski VA, Brunet A. Energy metabolism in adult neural stem cell fate. Prog Neurobiol. 2011;93(2):182–203. doi:10.1016/j.pneurobio.2010.10.007

55. Ojeda L, Gao J, Hooten KG, et al. Critical Role of PI3K/Akt/GSK3β in Motoneuron Specification from Human Neural Stem Cells in Response to FGF2 and EGF. Sham MH, ed. PLoS One. 2011;6(8):e23414. doi:10.1371/journal.pone.0023414

56. Wang XQ, Lo CM, Chen L, Ngan ESW, Xu A, Poon RYC. CDK1-PDK1-PI3K/Akt signaling pathway regulates embryonic and induced pluripotency. Cell Death Differ. 2017;24(1):38–48. doi:10.1038/cdd.2016.84

57. Marques L, Thorsteinsdóttir S. Dynamics of Akt activation during mouse embryo development: Distinct subcellular patterns distinguish proliferating versus differentiating cells. Differentiation. 2013;86(1-2):48–56. doi:10.1016/j.diff.2013.07.001

58. Liang Q, Luo Z, Zeng J, et al. Zika Virus NS4A and NS4B Proteins Deregulate Akt-mTOR Signaling in Human Fetal Neural Stem Cells to Inhibit Neurogenesis and Induce Autophagy. Cell Stem Cell. 2016;19(5):663–671. doi:10.1016/j.stem.2016.07.019

59. Li H, Saucedo-Cuevas L, Shresta S, Gleeson JG. The Neurobiology of Zika Virus. Neuron. 2016;92(5):949–958. doi:10.1016/j.neuron.2016.11.031

60. Schroering AG, Edelbrock MA, Richards TJ, Williams KJ. The cell cycle and DNA mismatch repair. Exp Cell Res. 2007;313(2):292–304. doi:10.1016/j.yexcr.2006.10.018

61. Tichy ED, Stambrook PJ. DNA repair in murine embryonic stem cells and differentiated cells. Exp Cell Res. 2008;314(9):1929–1936. doi:10.1016/j.yexcr.2008.02.007

62. Zhang H, Richards B, Wilson T, et al. Apoptosis Induced by Overexpression of HMSH2 or HMLH1 1. Vol 59.; 1999.

63. Daikoku T, Kudoh A, Sugaya Y, et al. Postreplicative mismatch repair factors are recruited to Epstein-Barr virus replication compartments. J Biol Chem. 2006;281(16):11422–11430. doi:10.1074/jbc.M510314200

64. Mohni KN, Mastrocola AS, Bai P, Weller SK, Heinen CD. DNA Mismatch Repair Proteins Are Required for Efficient Herpes Simplex Virus 1 Replication. J Virol. 2011;85(23):12241–12253. doi:10.1128/jvi.05487-11

65. Lei M. The MCM Complex: Its Role in DNA Replication and Implications for Cancer Therapy. Curr Cancer Drug Targets. 2005;5(5):365–380. doi:10.2174/1568009054629654

66. Ryu S, Holzschuh J, Erhardt S, Ettl A-K, Driever W. Depletion of Minichromosome Maintenance Protein 5 in the Zebrafish Retina Causes Cell-Cycle Defect and Apoptosis. Vol 102.; 2005. www.pnas.orgcgidoi10.1073pnas.0506187102. Accessed July 4, 2020.

67. Lee KY, Im JS, Shibata E, et al. MCM8-9 complex promotes resection of double-strand break ends by MRE11-RAD50-NBS1 complex. Nat Commun. 2015;6(1):1–12. doi:10.1038/ncomms8744

68. Wang Y, Cortez D, Yazdi P, Neff N, Elledge SJ, Qin J. BASC, a super complex of BRCA1-associated proteins involved in the recognition and repair of aberrant DNA structures. Genes Dev. 2000;14(8):927–939. doi:10.1101/gad.14.8.927

69. Balasubramanian N, Bai P, Buchek G, Korza G, Weller SK. Physical Interaction between the Herpes Simplex Virus Type 1 Exonuclease, UL12, and the DNA Double-Strand Break-Sensing MRN Complex. J Virol. 2010;84(24):12504–12514. doi:10.1128/jvi.01506-10

70. (PDF) Role of MRE11 in Cell Proliferation, Tumor Invasion, and DNA Repair in Breast Cancer. https://www.researchgate.net/publication/230723928_Role_of_MRE11_in_Cell_Proliferation_Tumor_Invasion_and_DNA_Repair_in_Breast_Cancer. Accessed July 4, 2020.

71. Damiola F, Pertesi M, Oliver J, et al. Rare key functional domain missense substitutions in MRE11A, RAD50, and NBN contribute to breast cancer susceptibility: Results from a Breast Cancer Family Registry case-control mutation-screening study. Breast Cancer Res. 2014;16(3). doi:10.1186/bcr3669

72. Bellacosa A. Developmental disease and cancer: Biological and clinical overlaps. Am J Med Genet Part A. 2013;161(11):2788–2796. doi:10.1002/ajmg.a.36267

73. Polymalformative genetic syndrome with increased risk of developing cancer (Concept Id: CN200541) - MedGen - NCBI. https://www.ncbi.nlm.nih.gov/medgen/797542. Accessed July 4, 2020.

74. Schiffman JD, Geller JI, Mundt E, Means A, Means L, Means V. Update on pediatric cancer predisposition syndromes. In: Pediatric Blood and Cancer. Vol 60. Pediatr Blood Cancer; 2013:1247–1252. doi:10.1002/pbc.24555

